# Benchmarking Open-Source Vision-Language Models for Brain Metastasis Assessment on Single-Slice Contrast-Enhanced MRI

**DOI:** 10.64898/2026.08.24.26361169

**Authors:** Junsung Kim, Bum-Soo Kim, Ji Su Ko, Jaejune Dong, Seo Yeon Youn, Jinhee Jang, Kook-Jin Ahn

## Abstract

**Purpose:** Open-source vision-language models (VLMs) can be locally deployed without external internet access. This study aimed to evaluate the slice-level lesion-detection performance of multiple general-purpose and medical-purpose open-source VLMs for brain metastases on contrast-enhanced (CE) MRI, and to assess their accuracy in characterizing detailed imaging features on lesion-positive images.

**Materials and Methods:** One hundred lesion-positive axial CE T1-weighted images and 100 matched lesion-negative images from 100 patients were analyzed using eight evaluable VLMs: five general-purpose VLMs–InternVL3-8B, Qwen2.5-VL-7B-Instruct, MiniCPM-V-4.5, DeepSeek-VL2-tiny, and Phi-3.5-vision-instruct–and three medical-purpose VLMs–MedGemma-4B-it, LLaVA-Med v1.5, and HuatuoGPT-Vision-7B. Lesion-detection performance was assessed using sensitivity, specificity, and balanced accuracy. On lesion-positive images, accuracy was evaluated for lesion count, laterality, anatomic location, enhancement pattern, necrosis, and mass effect. Balanced accuracy was reported with patient-level cluster bootstrap 95% confidence intervals (CIs). Models were compared using the Friedman test and paired patient-level permutation tests with Benjamini-Hochberg correction. Secondary outcomes were compared with majority-class baselines using exact McNemar tests. A revised prompt was tested post hoc.

**Results:** The median age of the study patients was 65 years (IQR, 59.0–70.0), and 58 patients were male (58.0%). MiniCPM-V-4.5 showed significantly higher balanced accuracy (80.5%; 95% CI, 75.0–85.5) than all other models (FDR-adjusted P < 0.001). Seven of the eight models showed predicted-positive rates of 81–100%, and 70.8% (17/24) of responses to synthetic control images without anatomic content were classified as lesion-positive. With the revised prompt, balanced accuracy increased from 59.5% to 77.0% in HuatuoGPT-Vision-7B and from 51.5% to 68.5% in InternVL3-8B. No model significantly outperformed the majority-class baseline for enhancement pattern or for mass effect.

**Conclusion:** This slice-level technical benchmark demonstrated variable lesion-detection performance of open-source VLMs for brain metastasis in an image-only setting, with most models showing a strong positive-response tendency.

## Introduction

Vision-language models (VLMs) are multimodal large language models that integrate visual and textual information [1]. Applications of VLMs to lesion detection and image interpretation have attracted increasing interest and have been rapidly evolving in radiology artificial intelligence (AI) [2]. Previous studies have consistently reported that VLM performance is substantially improved when text input, such as clinical information or radiologist-defined image descriptions are provided in addition to images [3,4]. Large language models and VLMs have also been shown to be effective in performing straightforward radiologic tasks such as MR sequence selection and imaging modality identification [5,6]. In addition, VLMs may support radiologists in clinical workflows [7,8]. However, their clinical utility remains limited by insufficient diagnostic accuracy, inconsistent image interpretation, and the possibility of hallucinations [9–15].

Radiologic images contain sensitive patient information, and the integration of AI into radiology workflows poses privacy and cybersecurity concerns [16–18]. At our institution, hospital computers are strictly isolated from external internet access due to concerns regarding patient privacy and data security. Therefore, locally deployable open-source VLMs may represent a practical alternative to cloud-based models (e.g., ChatGPT and Claude) in such environments.

Several benchmark studies have evaluated open-source VLMs, with or without comparisons with cloud-based VLMs, across various clinical settings, including brain tumor diagnosis, emergency and critical care, and neuroradiologic image interpretation [19–21]. These studies have consistently demonstrated insufficient diagnostic performance when visual image data were directly incorporated. Another multicenter benchmark and reader study has shown the potential of open-source large language models for generating diagnostic impressions from brain MRI reports, which may suggest stronger performance with text-based inputs than with visual inputs [22]. In addition, a recent study evaluated two cloud-based VLMs for detecting brain metastasis using both image and clinical context, and reported perfect sensitivity but very low specificity [23]. However, to our knowledge, no previous study has compared multiple contemporary, locally deployable open-source VLMs for detecting brain metastasis using a standardized image-only dataset. Therefore, the purpose of this study was to evaluate the slice-level lesion-detection performance of multiple general-purpose and medical-purpose open-source VLMs for brain metastases on contrast-enhanced (CE) MRI, and to assess their accuracy in characterizing detailed imaging features on lesion-positive images.

## Materials and Methods

This retrospective study was approved by the Institutional Review Board of Seoul St. Mary’s Hospital (IRB No. 2026-1376-0001), with a waiver of informed consent. Brain MR images were obtained from the AI Hub project (www.aihub.or.kr), supported by the Ministry of Science and ICT and the National Information Society Agency, Republic of Korea.

### Patient and study design

This study used single-slice axial CE T1-weighted images (T1WI) from the AI Hub brain metastasis dataset. The original dataset was collected at Gachon University Gil Medical Center. Among the 989 cases in the AI Hub dataset, 100 consecutive patients were evaluated using nine VLMs (eight evaluable). The first 100 available cases (from GMC-0001 to GMC-0112) were included, as some identification numbers were not assigned in the original dataset (n=10) and clinical data were missing in two cases (n=2). No additional inclusion or exclusion criteria were applied. The MR acquisition period of the study patients was from September 2019 to December 2020. The study workflow is summarized in **Figure 1**. The baseline demographic characteristics of the 100 analyzed patients were compared with those of the remaining (not-analyzed) patients in the AI Hub dataset, and the results are presented in **Supplementary Table S1**.

**Figure 1.**
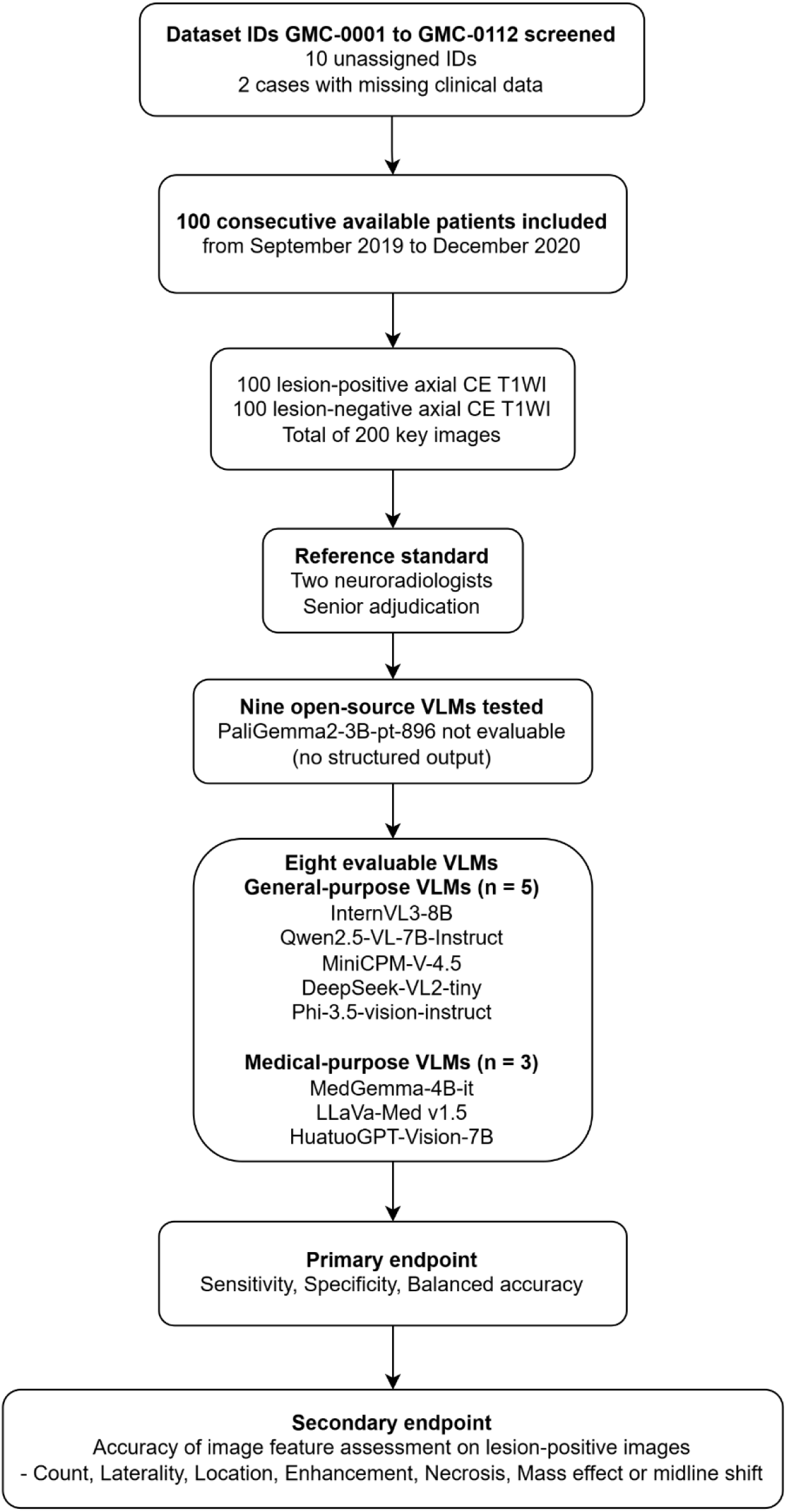
Study workflow.

### Computational environment and Vision-Language Models

All experiments were conducted in the AI Hub Secure Zone, an offline computing environment without external internet access. Online connection was temporarily permitted for the initial setup of the computing environment, including installation of required software packages and deployment of VLMs. All external network connections were disabled after the brain metastasis dataset was mounted to the virtual workstation.

The provided virtual workstation was equipped with a single NVIDIA Tesla V100-SXM2 GPU (32 GB VRAM) running CUDA 12.2 and Python 3.10. Considering the offline computing environment and available computational resources, open-source VLMs with 3–8 billion parameters that could be successfully executed within the secure environment were selected for analysis. Access to the AI Hub Secure Zone was available from June 18, 2026 to October 2, 2026, during which all experiments were performed.

The evaluated open-source VLMs consisted of 6 general-purpose VLMs and 3 medical-purpose VLMs. General-purpose VLMs included InternVL3-8B, Qwen2.5-VL-7B-Instruct, MiniCPM-V-4.5, DeepSeek-VL2-tiny, Phi-3.5-vision-instruct, and PaliGemma2-3B-pt-896. Medical-purpose VLMs were MedGemma-4B-it, LLaVA-Med v1.5, and HuatuoGPT-Vision-7B. All models were evaluated using deterministic decoding (do_sample=False or equivalent; maximum 512 new tokens) without stochastic sampling [24]. Exact model revisions are provided in **Supplementary Tables S2A and S2B**.

### Establishment of reference standard

The original brain metastasis dataset consisted only of DCM files. DCM files were converted to PNG files using the pydicom library after clipping pixel intensities at the 1st and 99th percentiles and scaling to 8 bits. The Python script used for converting DCM into PNG is provided in the **Supplementary Script 1**.

100 lesion-positive and 100 matched lesion-negative axial CE T1WI images were initially selected by one neuroradiologist (J.S.Kim, 6 years of experience in neuroradiology), and subsequently verified by a second neuroradiologist (J.S.Ko, 3 years of experience in neuroradiology). For lesion-positive images, the most conspicuous image was selected by J.S.Kim. Lesion-negative matched images were obtained from the same patients with confirmed brain metastasis, selected at anatomic levels different from the metastatic lesion. Lesion-negative matched images were obtained from three anatomical levels, including centrum semiovale (CS; n=32), basal ganglia (BG; n=34), and middle cerebellar peduncle (MCP; n=34). These levels were selected to provide representative supratentorial and infratentorial normal brain regions.

Reference standards for lesion count, laterality and lesion location were established by consensus of two neuroradiologists (J.S.Kim and J.S.Ko). The same readers independently classified: (1) enhancement pattern (homogeneous, heterogeneous, ring, or NA [not assessable]), (2) necrosis (yes/no), and (3) mass effect or midline shift (yes/no). Discrepancies were resolved by a senior neuroradiologist (B.S.K., 31 years of experience in neuroradiology) [25]. Reference standards were established from the imaging findings without access to model outputs during the final 100-case review, which was performed after a two-month washout period. The readers had previously been exposed to outputs from the initial 60-case analysis.

The reference lesion count was defined as the number of enhancing lesions on the selected single-slice key image. Lesions that were visible only on other slices were not included in the reference lesion count. Lesion counts were categorized into single versus multiple, and as predefined groups of 1, 2, 3, 4, 5, and ≥ 6 lesions. Model-generated lesion counts were compared with both reference categories.

Lesion location was recorded for all enhancing lesions on the key image and classified as supratentorial, infratentorial, or both (**Table 1**). For the secondary endpoint analyses, the largest enhancing lesion on the key image was used as the reference lesion for location (supratentorial vs. infratentorial), enhancement pattern, and necrosis. Necrosis was defined as a centrally hypoenhancing area within the enhancing lesion. In contrast, laterality, lesion count, and mass effect were assessed for the key image as a whole.

**Table 1.** Clinical and tumor characteristics of the study cohort.

| **Clinical variables (n=100)** | **n (%)** |
| --- | --- |
| **Age (years), median [IQR]** | 65.0 [59.0–70.0] |
| **Sex** |  |
| Male | 58 (58.0) |
| Female | 42 (42.0) |
| **Primary tumor site** |  |
| Lung | 63 (63.0) |
| Breast | 17 (17.0) |
| Colorectum | 9 (9.0) |
| Others | 11 (11.0) |
| **Tumor characteristics (n=100)** | **n (%)** |
| **Number of brain metastases (key image)** |  |
| Single (1) | 62 (62.0) |
| Multiple | 38 (38.0) |
| 2 | 14 (14.0) |
| 3 | 6 (6.0) |
| 4 | 4 (4.0) |
| 5 | 3 (3.0) |
| ≥6 | 11 (11.0) |
| **Laterality (key image)** |  |
| Right | 42 (42.0) |
| Left | 29 (29.0) |
| Bilateral | 29 (29.0) |
| **Location (all lesions on the key image)<sup>1</sup>** |  |
| Supratentorial | 72 (72.0) |
| Infratentorial | 22 (22.0) |
| Supratentorial and infratentorial | 6 (6.0) |
| **Enhancement (largest lesion)** |  |
| Ring enhancement | 21 (21.0) |
| Non-ring enhancement | 79 (79.0) |
| Homogeneous | 41 (41.0) |
| Heterogeneous | 38 (38.0) |
| **Necrosis (largest lesion), present** | 49 (49.0) |
| **Mass effect or midline shift (key image), present** | 13 (13.0) |
| **Lesion size (largest lesion), cm, median [IQR]** | 1.3 [0.7–2.2] |

<sup>1</sup>In the secondary analyses, the largest lesion was supratentorial in 75 and infratentorial in 25 patients Data are median [interquartile range] for age and size and number (%) otherwise.

MicroDicom Portable was used to view original DCM files. The lesion size was measured as a maximum length of the largest lesion on the key image. In addition, the converted PNG images were reviewed by a neuroradiologist (J.S.Kim) to cross-check the orientation of all key images.

Representative InternVL3-8B outputs for lesion-positive and lesion-negative key images are presented in **Figure 2**.

**Figure 2.**
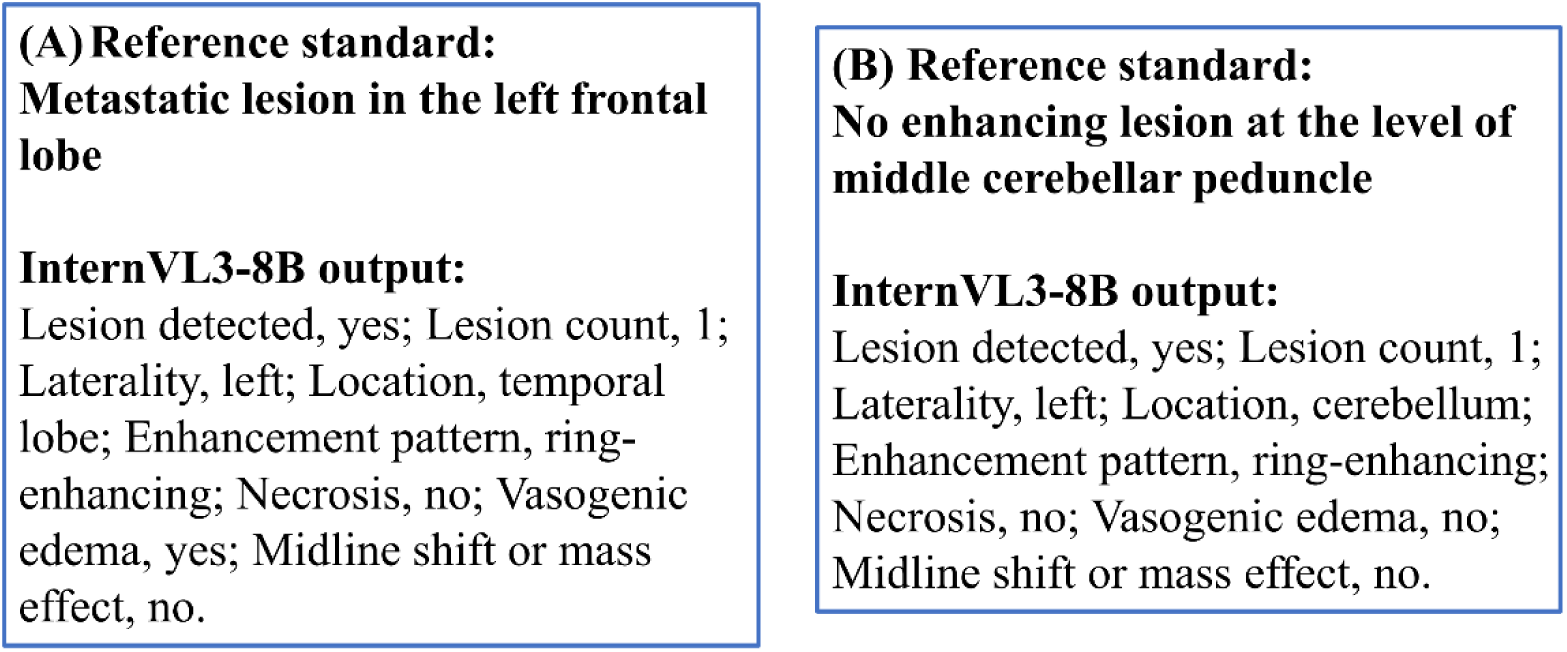
Representative InternVL3-8B outputs for lesion-positive and lesion-negative key images. (A) The reference standard indicated a metastatic lesion in the left frontal lobe, whereas InternVL3-8B incorrectly identified its location as a temporal lobe. (B) The reference standard indicated no enhancing lesion at the level of middle cerebellar peduncle, whereas InternVL3-8B generated a false-positive response. * Note: The AI Hub Secure Zone does not allow the export of source medical images under any circumstances. Therefore, only the corresponding text outputs are presented to illustrate the model response format.

### Outcome measures

The primary endpoint of the study was lesion-detection sensitivity, specificity, and balanced accuracy (i.e. mean of sensitivity and specificity) of open-source VLMs among lesion-positive and lesion-negative images. The secondary endpoint was the accuracy of imaging feature assessment on lesion-positive images. Imaging feature assessments included lesion count, laterality, anatomic location, enhancement pattern, necrosis and mass effect or midline shift.

### Prompt design

The prompt was fixed before any VLM output was reviewed to avoid tuning it to the evaluated data. The same prompt was independently applied to each of the nine VLMs to ensure a standardized comparison across models. The prompt requested lesion detection (yes or no), lesion count, lesion laterality, detailed anatomic location, enhancement pattern, and the presence of necrosis, vasogenic edema, and mass effect or midline shift. The full prompt is provided in **Supplementary Script 2A.**

Vasogenic edema is inherently difficult to assess on single-slice CE T1WI because it is evaluated on T2-weighted or FLAIR images; this item was therefore not analyzed. In addition, after reviewing the VLM outputs for the initial prompt, we found that DeepSeek-VL2-tiny repeated the item names instead of answering several items (items 5 to 8). As a post hoc analysis, a revised prompt that specified the answer format for each item and omitted the vasogenic edema item was run once for all models. The revised script after VLM output verification is described in **Supplementary Script 2B**. The initial prompt was retained for the primary analysis.

Separate Python scripts were used for each VLM. Each pipeline script loaded the PNG key image, read the prompt text file, loaded the model, and automatically converted the model-generated outputs into a structured CSV file. A representative inference and structured-output extraction pipeline using InternVL3-8B is provided in **Supplementary Script 3**. Parsing accuracy was verified by comparing the structured outputs with the raw data using the original prompt in 10 randomly selected images per model (80 in total).

### Control experiment

Three synthetic images without anatomical content were generated with Python, including a uniform black image (pixel value 0 in all channels), a uniform mid-gray image (pixel value 128), and a noise image with pixel values drawn independently from a uniform distribution over 0–255 in each RGB channel. Each image was saved as an 8-bit RGB PNG with the same matrix size as a representative control key image. The black, gray, and noise images were presented to each model with the original prompt.

### Secondary endpoint analysis

Model outputs for the secondary imaging feature assessment were mapped to predefined categories. Blank, not applicable, item-name echo, and out-of-category responses were considered invalid and scored as incorrect. The items were lesion count (single vs. multiple and exact count), laterality (right, left, or bilateral), location (supratentorial vs. infratentorial), enhancement pattern (ring enhancement vs. non-ring enhancement), necrosis, and mass effect or midline shift.

For each item, accuracy was compared with the majority-class baseline. Majority-class baseline was defined as accuracy of a classifier that always predicts the most frequent reference category. For the exact lesion count, macro-F1 was also calculated. For the binary items, sensitivity, specificity and balanced accuracy were measured, with multiple lesions, infratentorial location, ring enhancement, necrosis, and mass effect as the positive classes. Laterality was evaluated by accuracy only.

### Statistical analysis

A target sample size of 100 lesion-positive and 100 lesion-negative key images was based on the expected confidence interval (CI) half-width of 6–10 percentage points. Continuous variables were presented as medians with interquartile ranges (IQRs).

Lesion-detection performance was evaluated by sensitivity and specificity with 95% CIs calculated by the Wilson score method. Since some VLMs showed markedly imbalanced sensitivity and specificity, balanced accuracy was used for statistical comparison. Because each patient contributed one lesion-positive and one lesion-negative image, the 95% CI of balanced accuracy was estimated by a patient-level cluster bootstrap (10,000 resamples). Balanced accuracy across models was compared with the Friedman test, and pairwise differences with paired patient-level sign-flip permutation tests (50,000 permutations) and Benjamini-Hochberg (BH) correction for 28 comparisons. Specificity for the three anatomical levels (CS, BG, MCP) of the lesion-negative key images was calculated with 95% CI and compared for each model with Fisher’s exact test, with BH correction across models.

The secondary endpoints were analyzed among the 100 lesion-positive key images. The 95% CIs of accuracy, sensitivity, and specificity were calculated by the Wilson score method. The 95% CIs of balanced accuracy for binary items were estimated by a percentile bootstrap (2,000 resamples). For each item, the accuracy of each model was compared with that of the majority-class classifier using the exact McNemar test with BH correction across 48 comparisons (8 models × 6 items). The exact lesion count was reported with accuracy and macro-F1. As additional analyses, the secondary endpoints were also calculated among the images in which the model reported a lesion (conditional on detection) and separately for single- and multiple-lesion cases.

Sensitivity for supratentorial and infratentorial lesions for largest lesion on lesion-positive key image was compared with Fisher’s exact test. Predicted-positive rates were calculated for each model for the lesion-positive and lesion-negative images and for the control images. For the post hoc analysis of the revised prompt, balanced accuracy was analyzed in the same way, with BH correction for 21 pairwise comparisons among the seven evaluable models. Inter-reader agreement for the independent classification of enhancement pattern, necrosis, and mass effect or midline shift was assessed with Cohen’s kappa. Baseline characteristics of the analyzed and not-analyzed patients were compared with the Wilcoxon rank-sum test for age, the chi-square test with continuity correction for sex, and Fisher’s exact test (Monte Carlo simulation, 10,000 replicates) for primary tumor site. Analyses were performed with R version 4.6.1, and two-sided P < 0.05 (BH-adjusted P < 0.05 for multiple comparisons) was considered statistically significant.

## Results

### Patients and models

The analyzed patients did not differ significantly from the remaining patients in the AI Hub dataset in age, sex or primary tumor site (all P > 0.2; **Supplementary Table S1**). The median age of the study patients was 65 years (IQR, 59.0–70.0 years), and 58 patients were male (58.0%). The most common primary tumor site was lung (n=63, 63.0%), followed by breast (n=17, 17.0%) and colorectal (n=9, 9.0%). Brain metastases were in the supratentorial region (n=72, 72.0%), the infratentorial region (n=22, 22.0%), or both regions (n=6, 6.0%). Clinical and tumor characteristics of the study cohort are summarized in **Table 1**.

PaliGemma2-3B-pt-896 was not evaluable because it returned no structured response for any of the 200 key images with either the original or the revised prompt, or for any of the three control images with the original prompt. The remaining eight models (five general-purpose and three medical-purpose VLMs) were therefore included in the analysis. All parsed fields (structured CSV file) matched the raw outputs (80/80, verified by J.S.Kim).

### Control experiment

A total of 24 responses (three synthetic images for each of the eight evaluable models) were analyzed. Among them, 70.8% (17/24) were positive (black, 6/8; gray, 6/8; noise, 5/8). MiniCPM-V-4.5 reported “no” for all three images, whereas InternVL3-8B, Qwen2.5-VL-7B-Instruct, DeepSeek-VL2-tiny, and LLaVA-Med v1.5 reported “yes” for all three. The detailed model outputs are presented in **Supplementary Table S3**.

### Lesion-detection performance of open-source VLMs

Balanced accuracy varied significantly across the eight models (Friedman test, P < 0.001). MiniCPM-V-4.5 demonstrated the highest balanced accuracy of 80.5% (95% CI, 75.0–85.5), with a sensitivity of 79.0% (95% CI, 70.0–85.8) and a specificity of 82.0% (95% CI, 73.3–88.3) (**Table 2**, **Figure 3**). The balanced accuracy of MiniCPM-V-4.5 was significantly higher than that of all other models (balanced accuracy difference, 19.5–30.5 percentage points; all FDR-adjusted P < 0.001; **Table 3**).

**Table 2.** Per-image lesion-detection performance of vision-language models.

| VLM | Sensitivity, % [95% CI] | Specificity, % [95% CI] | Balanced accuracy, % [95% CI] | Predicted positive rate, % |
| --- | --- | --- | --- | --- |
| **General-purpose VLMs** |  |  |  |  |
| InternVL3-8B | 100.0 [96.3–100.0] | 3.0 [1.0–8.5] | 51.5 [50.0–53.5] | 98.5 |
| Qwen2.5-VL-7B-Instruct | 92.0 [85.0–95.9] | 30.0 [21.9–39.6] | 61.0 [56.0–66.0] | 81.0 |
| MiniCPM-V-4.5 | 79.0 [70.0–85.8] | 82.0 [73.3–88.3] | 80.5 [75.0–85.5] | 48.5 |
| DeepSeek-VL2-tiny | 100.0 [96.3–100.0] | 0.0 [0.0–3.7] | 50.0 [50.0–50.0] | 100.0 |
| Phi-3.5-vision-instruct | 100.0 [96.3–100.0] | 2.0 [0.6–7.0] | 51.0 [50.0–52.5] | 99.0 |
| **Medical-purpose VLMs** |  |  |  |  |
| MedGemma-4B-it | 100.0 [96.3–100.0] | 2.0 [0.6–7.0] | 51.0 [50.0–52.5] | 99.0 |
| LLaVA-Med v1.5 | 100.0 [96.3–100.0] | 0.0 [0.0–3.7] | 50.0 [50.0–50.0] | 100.0 |
| HuatuoGPT-Vision-7B | 98.0 [93.0–99.4] | 21.0 [14.2–30.0] | 59.5 [55.5–64.0] | 88.5 |

Data are based on 100 lesion-positive and 100 lesion-negative key images from 100 patients. Sensitivity and specificity are given with Wilson 95% confidence intervals; the 95% confidence interval of balanced accuracy was estimated with a patient-level cluster bootstrap (10,000 resamples). Models that classified all images as positive have no variability in balanced accuracy. PaliGemma2-3B-pt-896 returned no structured output and was not evaluable.

**Figure 3.**
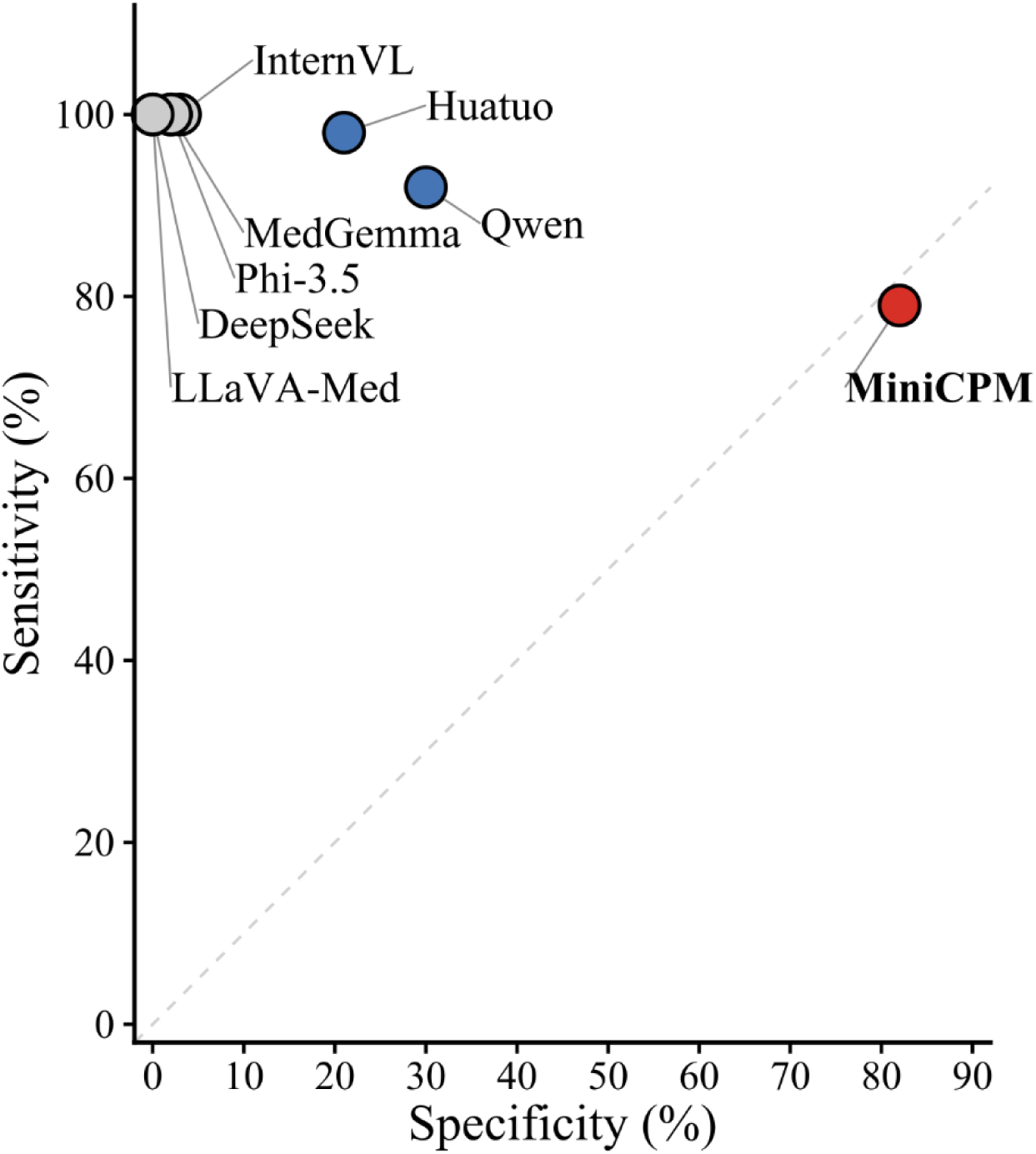
Model-wise sensitivity and specificity plot for brain metastasis detection. Each point represents the sensitivity and specificity of one VLM. The dashed diagonal line indicates equal sensitivity and specificity. MiniCPM-V-4.5 demonstrated the highest balanced accuracy, and is highlighted by a red circle. Qwen2.5-VL-7B-Instruct and HuatuoGPT-Vision-7B showed relatively intermediate balanced accuracy, and are highlighted by blue circles. * Abbreviations: InternVL, InternVL3-8B; Qwen, Qwen2.5-VL-7B-Instruct; MiniCPM, MiniCPM-V-4.5; MedGemma, MedGemma-4B-it; LLaVA-Med, LLaVA-Med v1.5; Huatuo, HuatuoGPT-Vision-7B; Phi-3.5, Phi-3.5-vision-instruct; DeepSeek, DeepSeek-VL2-tiny

**Table 3.**
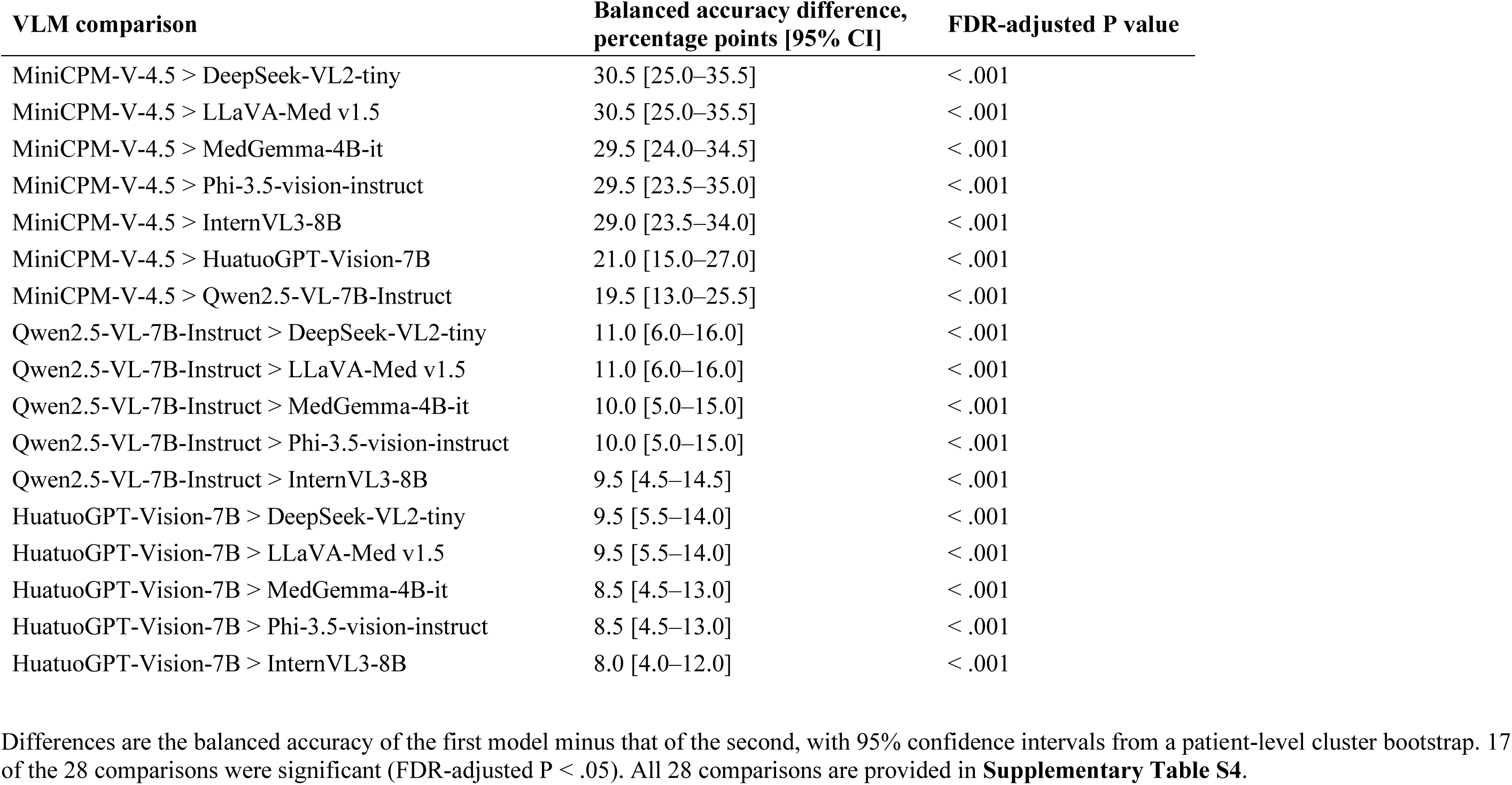
Significant pairwise comparisons of balanced accuracy among vision-language models.

| VLM comparison | Balanced accuracy difference, percentage points [95% CI] | FDR-adjusted P value |
| --- | --- | --- |
| MiniCPM-V-4.5 > DeepSeek-VL2-tiny | 30.5 [25.0–35.5] | < .001 |
| MiniCPM-V-4.5 > LLaVA-Med v1.5 | 30.5 [25.0–35.5] | < .001 |
| MiniCPM-V-4.5 > MedGemma-4B-it | 29.5 [24.0–34.5] | < .001 |
| MiniCPM-V-4.5 > Phi-3.5-vision-instruct | 29.5 [23.5–35.0] | < .001 |
| MiniCPM-V-4.5 > InternVL3-8B | 29.0 [23.5–34.0] | < .001 |
| MiniCPM-V-4.5 > HuatuoGPT-Vision-7B | 21.0 [15.0–27.0] | < .001 |
| MiniCPM-V-4.5 > Qwen2.5-VL-7B-Instruct | 19.5 [13.0–25.5] | < .001 |
| Qwen2.5-VL-7B-Instruct > DeepSeek-VL2-tiny | 11.0 [6.0–16.0] | < .001 |
| Qwen2.5-VL-7B-Instruct > LLaVA-Med v1.5 | 11.0 [6.0–16.0] | < .001 |
| Qwen2.5-VL-7B-Instruct > MedGemma-4B-it | 10.0 [5.0–15.0] | < .001 |
| Qwen2.5-VL-7B-Instruct > Phi-3.5-vision-instruct | 10.0 [5.0–15.0] | < .001 |
| Qwen2.5-VL-7B-Instruct > InternVL3-8B | 9.5 [4.5–14.5] | < .001 |
| HuatuoGPT-Vision-7B > DeepSeek-VL2-tiny | 9.5 [5.5–14.0] | < .001 |
| HuatuoGPT-Vision-7B > LLaVA-Med v1.5 | 9.5 [5.5–14.0] | < .001 |
| HuatuoGPT-Vision-7B > MedGemma-4B-it | 8.5 [4.5–13.0] | < .001 |
| HuatuoGPT-Vision-7B > Phi-3.5-vision-instruct | 8.5 [4.5–13.0] | < .001 |
| HuatuoGPT-Vision-7B > InternVL3-8B | 8.0 [4.0–12.0] | < .001 |

Qwen2.5-VL-7B-Instruct and HuatuoGPT-Vision-7B formed an intermediate group, with balanced accuracies of 61.0% (95% CI, 56.0–66.0) and 59.5% (95% CI, 55.5–64.0), respectively. These two models did not differ from each other but were significantly higher than the remaining five models.

InternVL3-8B, DeepSeek-VL2-tiny, Phi-3.5-vision-instruct, MedGemma-4B-it, and LLaVA-Med v1.5 classified 98.5–100.0% of all key images as positive (**Table 2**), with a sensitivity of 100.0% and a specificity of 0.0–3.0%. Complete pairwise comparison results are provided in the **Supplementary Table S4.**

### Specificity of lesion-negative images

LLaVA-Med v1.5 and DeepSeek-VL2-tiny have classified all lesion-positive and lesion-negative images as “positive” (i.e. specificity of 0%); therefore, the differences across levels for these models were not available.

Specificity was substantially different according to different anatomical levels. At the MCP level, all models except MiniCPM-V-4.5 (70.6%, 24/34) classified every lesion-negative slice as positive (specificity, 0%). Specificity was highest at the CS level for all models except DeepSeek-VL2-tiny and LLaVA-Med v1.5 (MiniCPM-V-4.5, 100%, 32/32; Qwen2.5-VL-7B-Instruct, 75%, 24/32; HuatuoGPT-Vision-7B, 50%, 16/32). The differences across anatomical levels were statistically significant for InternVL3-8B, Qwen2.5-VL-7B-Instruct, MiniCPM-V-4.5, and HuatuoGPT-Vision-7B (**Supplementary Table S5**).

### Post hoc analysis of models with revised prompt

LLaVA-Med v1.5 returned no output for any image and was not evaluable. Therefore, seven models were included in the post hoc sensitivity analysis (**Supplementary Tables S6 and S7**). MiniCPM-V-4.5 again showed the highest balanced accuracy (82.5%), followed by HuatuoGPT-Vision-7B (77.0%; not significantly different, FDR-adjusted P = 0.141) and InternVL3-8B (68.5%). DeepSeek-VL2-tiny and Phi-3.5-vision-instruct classified all images as positive and negative, respectively (balanced accuracy, 50.0%).

### Imaging feature assessment on lesion-positive images

The largest lesion was supratentorial in 75 and infratentorial in 25 patients, which is consistent with the known predominance of supratentorial brain metastases [26]. Sensitivity did not differ significantly between supratentorial and infratentorial lesions in the three models that did not classify all images as positive; MiniCPM-V-4.5 (77.3% [58/75] vs. 84.0% [21/25], P = 0.580), Qwen2.5-VL-7B-Instruct (89.3% [67/75] vs. 100% [25/25], P = 0.195), and HuatuoGPT-Vision-7B (97.3% [73/75] vs. 100% [25/25], P = 1.000). The remaining five models detected all lesions as positive in both compartments. The lobar region of the largest lesion is described in **Supplementary Table S8**.

Majority-class baselines for lesion count, laterality, location, enhancement, necrosis, mass effect or midline shift were 62%, 42%, 75%, 79%, 51%, and 87%, respectively. Of the 48 comparisons with the majority-class baseline, accuracy was significantly higher in 11 and lower in 12. HuatuoGPT-Vision-7B exceeded the baseline for lesion count, laterality, and location. MedGemma-4B-it exceeded the baseline for lesion count, laterality, and necrosis. Accuracy was lower than the baseline for enhancement in all models except for Phi-3.5-vision-instruct. No model exceeded the 87% baseline for midline shift or mass effect (**Table 4**).

**Table 4.** Accuracy of vision-language models for imaging feature assessment among lesion-positive key images (n=100)

| **Model** | **Lesion count**<br>**single vs.**<br>**multiple** | **Laterality** | **Location**<br>**Supra- vs.**<br>**Infratentorial** | **Enhancement**<br>**Ring vs. Non-ring** | **Necrosis** | **Mass effect or**<br>**midline shift** |
| --- | --- | --- | --- | --- | --- | --- |
| Majority-class baseline<sup>1</sup> | 62 (single) | 42 (Right) | 75 (Supratentorial) | 79 (Non-ring<br>enhancement) | 51 (No) | 87 (No) |
| InternVL3-8B | 71<sup>†</sup> | 30 | 88<sup>†</sup> | 34<sup>‡</sup> | 56 | 87 |
| Qwen2.5-VL-7B-Instruct | 61 | 42 | 86<sup>†</sup> | 21<sup>‡</sup> | 57 | 39<sup>‡</sup> |
| MiniCPM-V-4.5 | 63 | 40 | 72 | 36<sup>‡</sup> | 49 | 63<sup>‡</sup> |
| DeepSeek-VL2-tiny<sup>2</sup> | 70<sup>†</sup> | 42 | 0<sup>‡</sup> | 0<sup>‡</sup> | 0<sup>‡</sup> | 0<sup>‡</sup> |
| Phi-3.5-vision-instruct | 62 | 42 | 64 | 79 | 48 | 87 |
| MedGemma-4B-it | 76<sup>†</sup> | 64<sup>†</sup> | 79 | 57<sup>‡</sup> | 68<sup>†</sup> | 85 |
| LLaVA-Med v1.5 | 50 | 29 | 83<sup>†</sup> | 42<sup>‡</sup> | 49 | 87 |
| HuatuoGPT-Vision-7B | 78<sup>†</sup> | 79<sup>†</sup> | 94<sup>†</sup> | 26<sup>‡</sup> | 52 | 87 |

Data are accuracy (%).
<sup>1</sup>Accuracy of a classifier that always predicts the most frequent reference category (in parentheses). Accuracy is the percentage of the 100 lesion-positive key images correctly classified; images in which the model did not report a lesion and invalid responses were counted as incorrect.
<sup>2</sup>All responses were invalid for location, enhancement, necrosis, mass effect or midline shift (item names repeated, or "left"/"right" instead of an anatomical location) and were counted as incorrect (accuracy, 0%).
<sup>†</sup>Significantly higher than the baseline (exact McNemar test, Benjamini–Hochberg-adjusted $P < 0.05$ for 48 comparisons)
<sup>‡</sup>Significantly lower than the baseline (exact McNemar test, Benjamini–Hochberg-adjusted $P < 0.05$ for 48 comparisons)

Balanced accuracy for the binary imaging features is shown in **Table 5** (sensitivity, specificity and counts in **Supplementary Table S9**). Balanced accuracy was highest for location (HuatuoGPT-Vision-7B, 93.3%; InternVL3-8B, 80.0%), for mass effect or midline shift (HuatuoGPT-Vision-7B, 86.0%), and for lesion count (single vs. multiple; HuatuoGPT-Vision-7B, 72.1%; MedGemma-4B-it, 71.5%).

**Table 5.** Balanced accuracy of vision-language models for binary imaging features among lesion-positive key images (n=100)

| **Model** | **Lesion count**<br>**single vs. multiple** | **Location**<br>**Supra- vs. Infratentorial** | **Enhancement**<br>**Ring vs. Non-ring** | **Necrosis** | **Mass effect or**<br>**midline shift** |
| --- | --- | --- | --- | --- | --- |
| InternVL3-8B | 61.8 [55.3–69.2] | 80.0 [70.0–89.3] | 58.2 [54.3–62.6] | 55.2 [49.8–60.9] | 50.0 [50.0–50.0] |
| Qwen2.5-VL-7B-Instruct | 52.2 [45.6–59.5] | 76.0 [65.4–86.2] | 50.0 [50.0–50.0] | 57.8 [53.2–63.0] | 58.4 [46.5–69.0] |
| MiniCPM-V-4.5 | 59.0 [49.0–68.3] | 74.7 [64.4–84.0] | 57.7 [50.7–63.8] | 49.8 [43.5–56.1] | 75.5 [64.4–82.9] |
| DeepSeek-VL2-tiny | 60.5 [54.3–67.2] | 0.0 [0.0–0.0]<sup>†</sup> | 0.0 [0.0–0.0]<sup>†</sup> | 0.0 [0.0–0.0]<sup>†</sup> | 0.0 [0.0–0.0]<sup>†</sup> |
| Phi-3.5-vision-instruct | 50.0 [50.0–50.0] | 76.0 [70.5–81.5] | 50.0 [50.0–50.0] | 48.9 [45.4–52.3] | 50.0 [50.0–50.0] |
| MedGemma-4B-it | 71.5 [63.0–80.0] | 70.0 [59.3–80.6] | 64.0 [53.3–74.4] | 67.7 [58.9–76.1] | 84.8 [72.8–94.3] |
| LLaVA-Med v1.5 | 58.7 [52.5–65.2] | 66.0 [57.1–75.0] | 56.3 [45.6–65.7] | 50.0 [50.0–50.0] | 50.0 [50.0–50.0] |
| HuatuoGPT-Vision-7B | 72.1 [63.7–80.3] | 93.3 [86.5–98.4] | 53.2 [50.6–56.1] | 52.9 [49.0–57.1] | 86.0 [74.1–95.2] |

Balanced accuracy is shown only for binary items; laterality (three categories) is reported as accuracy in Table 4.
<sup>†</sup>All responses for this item were invalid (item names repeated, or "left"/"right" instead of an anatomical location), so balanced accuracy is 0.

For exact lesion count, accuracy ranged from 27% to 66%, and macro-F1 ranged from 0.10 to 0.28, with MedGemma-4B-it being the highest for both (**Supplementary Table S10**). Even for MedGemma-4B-it, the 95% CI of accuracy included the 62% baseline. Conditional-on-detection analyses and analyses stratified by single vs. multiple lesion cases are provided in **Supplementary Table S11**.

### Inter-reader agreement

Inter-reader agreement before senior adjudication was performed for enhancement pattern, necrosis, and mass effect or midline shift. Inter-reader agreement was substantial to almost perfect for necrosis (κ= 0.820; 95% CI, 0.707–0.932) and mass effect or midline shift (κ= 0.765; 95% CI, 0.585–0.944). Agreement for enhancement was moderate for both the four-category classification (κ=0.591; 95% CI, 0.469–0.712) and binary classification (ring vs. non-ring, κ=0.533; 95% CI, 0.351–0.715).

## Discussion

The current slice-level technical benchmark demonstrated substantial heterogeneity in the lesion-detection performance of open-source VLMs for brain metastasis evaluation. MiniCPM-V-4.5 achieved the highest balanced accuracy (80.5%) among the eight evaluable open-source VLMs with 3–8B parameters. Seven of the eight models showed predicted-positive rates of 81–100%, indicating a marked positive-response tendency in most evaluated VLMs. A post hoc analysis using the revised prompt revealed substantial sensitivity to prompt format in several models. In addition, no single model consistently outperformed the majority-class baselines across imaging feature assessments.

Synthetic black, gray, and noise images were tested using the original prompt, yielding positive responses in 70.8% (17/24) of trials. MiniCPM-V-4.5 was the only model that responded “no” to lesion presence across all three control images, which was consistent with its better lesion-detection performance in the primary outcome.

Our study aligns with the previous study reporting high sensitivity but low specificity in detecting brain metastasis by VLMs [23]. While Nelles *et al.* provided clinical history together with the images, the contribution of the image could not be separated from that of the accompanying text. In our image-only setting, the positive bias persisted without clinical context, suggesting that this tendency may not be attributable solely to textual information. Our study also showed that MiniCPM-V-4.5 had the potential to distinguish lesion-positive from lesion-negative slices under simplified conditions. MiniCPM-V-4.5 is an 8B-parameter VLM that employs a unified 3D-Resampler to reduce the number of visual tokens for image encoding [27]. Whether this architecture contributed to its favorable performance in lesion detection cannot be determined from the present study and warrants further research.

Several studies have described suboptimal diagnostic accuracy of VLMs and large language models in image-only settings [3,4,10–12,21]. HuatuoGPT-Vision-7B showed the highest balanced accuracy for lesion location (93.3%), whereas MedGemma-4B-it showed relatively balanced performance for lesion count (71.5%), location (70.0%), and necrosis (67.7%). However, no model significantly outperformed the majority-class baseline for mass effect or midline shift, or for enhancement pattern. These findings imply that imaging feature characterization may require more fine-grained visual interpretation than binary lesion detection, which may partly explain the observed inconsistency and modest accuracy across imaging feature assessments. Differences in model design and training data may also contribute to the heterogeneous performance among models [28].

Previous studies have reported superior diagnostic performance of cloud-based VLMs compared with open-source VLMs [20,21,29]. In addition, many VLM studies have focused on cloud-based VLMs because of their accessibility and ease of use. However, some institutions operate in offline environment because of privacy or data-governance policies, and cloud-based VLMs may not be feasible in such circumstances; open-source VLMs may provide a practical alternative.

While MiniCPM-V-4.5 maintained the highest balanced accuracy using the revised prompt, balanced accuracy increased from 59.5% to 77.0% in HuatuoGPT-Vision-7B, and from 51.5% to 68.5% in InternVL3-8B. While LLaVA-Med v1.5 returned outputs with the original prompt, it returned no output for any image and was not evaluable for the revised prompt. These findings may reflect prompt-based sensitivity of some of the evaluated models.

This study has several limitations. First, only a single-slice axial CE T1WI per case was analyzed. This may have limited the accurate evaluation of imaging features, and the single-slice image setting itself has inherent limitations for clinical application. Further studies using larger numbers of image inputs are warranted. Second, unlike a previous study with brain metastasis [23], lesion-negative images were extracted from the patients with confirmed metastases; the performance reported in our study therefore reflects slice-level rather than patient-level performance. Third, cloud-based VLMs were not included in this study. Fourth, larger open-source VLMs, such as Google/Gemma-4-31B-it and RadFM, were not evaluated because they were not available in the current virtual workstation environment. Our findings are therefore restricted to the evaluated 3–8B parameter models. Fifth, brain metastasis mimics, such as glioblastoma or primary CNS lymphoma, were not included. Sixth, although deterministic decoding was used, the reported performance reflects the specific VLM versions and computational environment, which may vary across model revisions or software environments. Seventh, a curated, single-center dataset was used, which may limit the generalizability of our results. Eighth, model-specific prompt format was not evaluated. Finally, the performance of radiologists was not compared with that of open-source VLMs, as this benchmark focused on relative comparison across VLMs.

In conclusion, our findings demonstrated variable lesion-detection performance of open-source VLMs for brain metastasis in a slice-level, image-only setting. Among the eight evaluated models, MiniCPM-V-4.5 achieved the highest balanced accuracy (80.5%), whereas most other models revealed a strong positive-response tendency. Further studies in clinically realistic settings, including disease-negative patients, disease mimics, multi-slice, multi-sequence MRI examinations, and interaction with radiologists, are warranted to determine the clinical value of open-source VLMs.

### AI usage statement

Claude (Anthropic) was used to assist with the preparation of the Python scripts (**Supplementary Scripts 1 and 3**) and **Supplementary Tables**. It was also used to assist with writing R code for additional statistical analyses including patient-level cluster bootstrap confidence intervals, the Friedman test, paired patient-level sign-flip permutation tests with Benjamini-Hochberg (BH) correction, and comparisons with majority-class baselines for the secondary outcome analyses. All code, statistical results and table contents were reviewed and verified by the authors. In addition, Claude (Anthropic) and ChatGPT (OpenAI) were used for English language editing of the manuscript.

## Declarations

### Ethics approval

This study was approved by the Institutional Review Board of Seoul St. Mary’s Hospital (IRB No. 2026-1376-0001).

### Consent to participate

The requirement for written informed consent was waived by the Institutional Review Board due to the retrospective nature of the study.

### Data Availability

The dataset analyzed in this study was accessed through AI Hub platform after institutional review board approval and is subject to the data access policies of AI Hub.

### Author contributions

J.S.Kim contributed to the study conception and design, data collection, ground-truth assessment, data analysis and interpretation, and drafting of the manuscript. B.S.K. contributed to the study conception and design, supervision, interpretation of the results, and critical revision of the manuscript. J.S.Ko contributed to ground-truth assessment and critical revision of the manuscript. J.J.D., S.Y.Y., J.H.J., and K.J.A. contributed to revision of the manuscript. All authors read and approved the final manuscript.

## Acknowledgements

None.

