## Supplementary material for "Benchmarking Open-Source Vision-Language Models for Brain Metastasis Assessment on Single-Slice Contrast-Enhanced MRI"

**Supplementary Scripts legends**

**Supplementary Script 1. Python script for DICOM-to-PNG conversion**

```
from pathlib import Path
import numpy as np
import pydicom
from PIL import Image

SOURCE_ROOT = Path("/path/to/source_dicom_directory")
OUTPUT_ROOT = Path("/path/to/output_png_directory")

def normalize_pixels(pixels, photometric):
	pixels = pixels.astype(np.float32)

	low = np.percentile(pixels, 1)
	high = np.percentile(pixels, 99)

	if high > low:
		pixels = np.clip(pixels, low, high)
		pixels = (pixels - low) / (high - low)
	else:
		pixels = np.zeros_like(pixels)

	pixels = (pixels * 255).astype(np.uint8)

	if photometric == "MONOCHROME1":
		pixels = 255 - pixels

	return pixels

def export_patient(patient_dir, output_root):
	patient_id = patient_dir.name
```

```
    out_dir =    output_root / patient_id / "png"
    out_dir.mkdir(parents=True, exist_ok=True)

    dicom_files = sorted(patient_dir.glob("*.dcm"))

    if not dicom_files:
        return f"[SKIP] {patient_id}: no DICOM files"

    for index, dicom_file in enumerate(dicom_files):
        ds = pydicom.dcmread(dicom_file)
        pixels = ds.pixel_array

        if pixels.ndim != 2:
            return f"[SKIP] {patient_id}: not 2D single-frame DICOM"

        photometric = str(getattr(ds, "PhotometricInterpretation", "MONOCHROME2"))
        pixels = normalize_pixels(pixels, photometric)

        image = Image.fromarray(pixels).convert("RGB")
        png_path = out_dir / f"frame_{index:03d}.png"
        image.save(png_path)

    return f"[OK] {patient_id}: {len(dicom_files)} DICOMS -> {out_dir}"

def main():
    OUTPUT_ROOT.mkdir(parents=True, exist_ok=True)

    patient_dirs = sorted([p for p in SOURCE_ROOT.iterdir() if p.is_dir()])

    print(f"Source root : {SOURCE_ROOT}")
    print(f"Output root : {OUTPUT_ROOT}")
    print(f"Patients found: {len(patient_dirs)}")
    print("=" *80)
```

```
    log_lines = []

    for patient_dir in patient_dirs:
        try:
            result = export_patient(patient_dir, OUTPUT_ROOT)
        except Exception as e:
            result = f"[ERROR] {patient_dir.name}: {e}"

        print(result)
        log_lines.append(result)

    log_path = OUTPUT_ROOT / "batch_export_log.txt"
    with open(log_path, "w", encoding="utf-8") as f:
        f.write("\n".join(log_lines))

    print("=" *80)
    print(f"Done. Log saved to: {log_path}")

if __name__ == "__main__":
    main()
```

**Supplementary Script 2. Prompt used in the study**

**Supplementary Script 2A. Original prompt**

Review this contrast-enhanced brain MRI image and answer briefly:

1. Enhancing lesion present: yes/no
2. Number of visible enhancing lesions
3. Laterality
4. Approximate anatomic location
5. Enhancement pattern
6. Central nonenhancing necrosis
7. Vasogenic edema
8. Visible mass effect or midline shift

Do not infer findings from sequences that are not provided.

Return only the eight requested items.

Do not include a disclaimer or any additional explanation.

**Supplementary Script 2B. Revised script after model output verification**

Review this contrast-enhanced brain MRI image and answer briefly:

1. Enhancing lesion present: yes/no
2. Number of visible enhancing lesions: 0, 1, 2, …
3. Laterality: left/right/bilateral
4. Approximate anatomic location:
5. Enhancement pattern:
6. Central nonenhancing necrosis: yes/no
7. Visible mass effect or midline shift: yes/no

If no lesion is visible, answer no, 0 then N/A for items 3-7.

Do not infer findings from sequences that are not provided.

Return only these items, with no disclaimer or explanation.

**Supplementary Script 3. Representative inference and structured-output extraction pipeline using InternVL3-8B.**

```
import os, csv, glob, re
import torch
from PIL import Image
from transformers import AutoModelForImageTextToText, AutoProcessor

os.environ["HF_HUB_OFFLINE"] = "1"
os.environ["TRANSFORMERS_OFFLINE"] = "1"

KEY_IMAGE_BASE = "/path/to/key_images"
PROMPT_FILE = "/path/to/prompts_brain_mri.txt"
OUTPUT_DIR = "/path/to/output/internvl_txt"
SUMMARY_CSV = "/path/to/output/internvl_structured_summary.csv"
MODEL = "/path/to/models/InternVL3-8B-hf"
MODEL_TAG = "internvl"

os.makedirs(OUTPUT_DIR, exist_ok=True)
os.makedirs(os.path.dirname(SUMMARY_CSV), exist_ok=True)
with open(PROMPT_FILE, encoding="utf-8") as f:
    prompt_text = f.read().strip()

def parse_output(text):
    d = {"LesionDetected":"","LesionCount":"","Laterality":"","Location":"",

"EnhancementPattern":"","Necrosis":"","VasogenicEdema":"","MassEffectOrMidlineShift":""}
    lines = [l.strip() for l in text.splitlines() if l.strip()]
    items = {}
    for l in lines:
        m = re.match(r'^\s*(\d+)[\.\)]\s*(.+)$', l)
        if m:
            items[int(m.group(1))] = m.group(2).strip()
    mapping = {1:"LesionDetected",2:"LesionCount",3:"Laterality",4:"Location",
```

```
5:"EnhancementPattern",6:"Necrosis",7:"VasogenicEdema",8:"MassEffectOrMidlineShift"}
    for num, key in mapping.items():
        if num in items:
            val = re.sub(r'^[A-Za-z ]+:\s*', '', items[num])
            d[key] = val
    return d

tasks = []
for case_dir in sorted(glob.glob(os.path.join(KEY_IMAGE_BASE, "GMC-*"))):
    case = os.path.basename(case_dir)
    for png in sorted(glob.glob(os.path.join(case_dir, "*.png"))):
        frame = os.path.splitext(os.path.basename(png))[0]
        tasks.append((case, frame, png))
print(f"Total images: {len(tasks)}")

print("Loading InternVL3-8B-hf...")
model = AutoModelForImageTextToText.from_pretrained(
    MODEL,
    torch_dtype=torch.float16,
    device_map="cuda:0",
    local_files_only=True,
    attn_implementation="sdpa",
).eval()
processor = AutoProcessor.from_pretrained(MODEL, local_files_only=True)
print("Model loaded.\n")

rows = []
for i, (case, frame, png) in enumerate(tasks, 1):
    image = Image.open(png).convert("RGB")
    messages = [{"role":"user","content":[
        {"type":"image","image":image},{"type":"text","text":prompt_text}]}]
    inputs = processor.apply_chat_template(messages, add_generation_prompt=True,
        tokenize=True, return_dict=True, return_tensors="pt")
```

```
        inputs = {k: (v.to("cuda:0") if isinstance(v, torch.Tensor) else v)
                  for k, v in inputs.items()}
        for k, v in inputs.items():
            if isinstance(v, torch.Tensor) and v.is_floating_point():
                inputs[k] = v.to(torch.float16)
        input_len = inputs["input_ids"].shape[-1]
        with torch.inference_mode():
            gen = model.generate(**inputs, max_new_tokens=512, do_sample=False)
            gen = gen[0][input_len:]
        out = processor.decode(gen, skip_special_tokens=True).strip()

        fname = f"{case}_{frame}__{MODEL_TAG}.txt"
        txt_path = os.path.join(OUTPUT_DIR, fname)
        with open(txt_path, "w", encoding="utf-8") as tf:
            tf.write(out)

        parsed = parse_output(out)
        rows.append({"Case":case, "File":fname, **parsed, "OriginalTxtPath":txt_path})
        print(f"   [{i}/{len(tasks)}] {case} {frame} -> {parsed['LesionDetected']}")

cols = ["Case","File","LesionDetected","LesionCount","Laterality","Location",

"EnhancementPattern","Necrosis","VasogenicEdema","MassEffectOrMidlineShift","OriginalTxtPath"
]
with open(SUMMARY_CSV, "w", newline="", encoding="utf-8") as f:
    w = csv.DictWriter(f, fieldnames=cols)
    w.writeheader()
    w.writerows(rows)

print(f"\nSaved summary: {SUMMARY_CSV}")
print(f"Total: {len(rows)} rows")
```

**Supplementary Table legends**

**Supplementary Table S1. Baseline characteristics of the analyzed patients compared with the remaining patients in the AI Hub brain metastasis dataset**

| | Analyzed (n=100) | Not analyzed (n=889) | Total (n=989) | P value |
|---|---|---|---|---|
| **Age** | 65.0 [59.0–70.0] | 65.0 [58.0–73.0] | 65.0 [58.0–73.0] | 0.844 |
| **Sex** | | | | 0.420 |
| F | 42 (42.0%) | 416 (46.8%) | 458 (46.3%) | |
| M | 58 (58.0%) | 473 (53.2%) | 531 (53.7%) | |
| **Primary tumor site** | | | | 0.236 |
| Lung | 63 (63.0%) | 594 (66.8%) | 657 (66.4%) | |
| Breast | 17 (17.0%) | 153 (17.2%) | 170 (17.2%) | |
| Colorectal | 9 (9.0 %) | 34 (3.8%) | 43 (4.3%) | |
| Others | 11 (11.0 %) | 102 (11.5%) | 113 (11.4%) | |
| Unknown | 0 (0.0 %) | 6 (0.7%) | 6 (0.6%) | |

Data are median [interquartile range] for age and number (%) for categorical variables. P values were obtained using the Wilcoxon rank-sum test for age, the chi-square test with continuity correction for sex, and the Fisher exact test for primary tumor site. "Others" comprised the biliary system, bladder, liver, nasal cavity, paranasal sinuses, urinary tract, pancreas, stomach, and uterus. "Unknown" indicates that the primary tumor site was not identified.

### Supplementary Table S2. Exact model revisions used in the study

#### Supplementary Table S2A. Evaluated open-source VLMs: Hugging Face repositories, exact versions (commit hashes), Transformers versions, and numerical precision

| Model | Category | Hugging Face repository | Commit hash | Transformers version | Precision |
|---|---|---|---|---|---|
| InternVL3-8B | General | OpenGVLab/InternVL3-8B-hf | 259a3b64a14623c0ec91a045cb43f7c5af5fa6af | 4.53.2 | float16 |
| Qwen2.5-VL-7B-Instruct | General | Qwen/Qwen2.5-VL-7B-Instruct | cc594898137f460bfe9f0759e9844b3ce807cfb5 | 4.53.2 | bfloat16 |
| MiniCPM-V-4.5 | General | openbmb/MiniCPM-V-4_5 | daef484c35ec93210ec93c5e901f8f3e9b78ee34 | 4.51.0 | bfloat16 |
| DeepSeek-VL2-tiny | General | deepseek-ai/deepseek-vl2-tiny | 66c54660eae7e90c9ba259bfdf92d07d6e3ce8aa | 4.38.2 | float16 |
| Phi-3.5-vision-instruct | General | microsoft/Phi-3.5-vision-instruct | 12b77fb40b63a2c73c68243d3f767aab688a1b2a | 4.43.0 | bfloat16 |
| PaliGemma2-3B-pt-896 | General | google/paligemma2-3b-pt-896 | 5a5194a3f819fc969b931c42f417c3194f5b6e4a | 4.53.2 | float16 |
| MedGemma-4B-it | Medical | google/medgemma-4b-it | 290cda5eeccbee130f987c4ad74a59ae6f196408 | 4.51.0 | bfloat16 |
| LLaVA-Med v1.5[1] | Medical | chaoyinshe/llava-med-v1.5-mistral-7b-hf | 627be53734c667cbb1669608dac747a4485a22d7 | 4.53.2 | float16 |
| HuatuoGPT-Vision-7B | Medical | FreedomIntelligence/HuatuoGPT-Vision-7B-hf | a092ca6536fe74015770f603e24b4e1538f0f2ac | 4.53.2 | float16 |
| **Model** | **Category** | **GitHub repository** | **Commit hash** | | |
| DeepSeek-VL2-tiny[2] | General | github.com/deepseek-ai/DeepSeek-VL2 | ef9f91e2b6426536b83294c11742c27be66361b1 | | |

[1]LLaVA-Med v1.5 is a Hugging Face-converted checkpoint derived from microsoft/llava-med-v1.5-mistral-7b.
[2]DeepSeek-VL2-tiny is not implemented in the Transformers library; its implementation code was installed from GitHub in addition to the model weights

**Supplementary Table S2B. Inference pipeline of each model**

| Model | Model class | Image loading | Image preprocessing | Prompt formatting | Generation function |
|---|---|---|---|---|---|
| InternVL3-8B | AutoModelForImageTextToText | PIL RGB object | Default processor settings | apply_chat_template() | generate() |
| Qwen2.5-VL-7B-Instruct | Qwen2_5_VLForConditionalGeneration | file path | min_pixels=256*28*28, max_pixels=1024*28*28 | apply_chat_template() | generate(), repetition_penalty=1.0 |
| MiniCPM-V-4.5 | AutoModel, AutoTokenizer (Remote code[1]; trust_remote_code=True) | PIL RGB object | No settings specified; default processing within the model's chat() method | Message list (user role; image and text) passed to chat(); chat template applied internally by chat() | chat() (method of the model's remote code); sampling = False, num_beams = 1, repetition_penalty = 1.0 |
| DeepSeek-VL2-tiny | AutoModelForCausalLM | PIL RGB object | Default DeepseekVLV2 Processor settings | Manually constructed conversation | prepare_inputs_embeds() (deepseek_vl2 package) followed by generate(); repetition_penalty = 1.0 |
| Phi-3.5-vision-instruct | AutoModelForCausalLM, AutoProcessor (Remote code; trust_remote_code=True) | PIL RGB object | num_crops=16 | apply_chat_template() | generate() |
| PaliGemma2-3B-pt-896 | PaliGemmaForConditionalGeneration, AutoProcessor | PIL RGB object | Default processor settings | No chat template; prompt passed as plain text | generate() |
| MedGemma-4B-it | AutoModelForImageTextToText, AutoProcessor | PIL RGB object | Default processor settings | apply_chat_template() | generate() |
| LLaVA-Med v1.5 | LlavaForConditionalGeneration, AutoProcessor | PIL RGB object | Default processor settings | apply_chat_template() | generate() |
| HuatuoGPT-Vision-7B | LlavaForConditionalGeneration, AutoProcessor | PIL RGB object | Default processor settings | Manually constructed Qwen2-style template with an <image> token (no chat template in the checkpoint) | generate() |

All models received the same PNG key image and text prompt. PNG files were generated from DICOM files without resizing (**Supplementary Script 1**) and were loaded from disk as 8-bit RGB bitmaps using PIL (Image.open().convert("RGB")); for Qwen2.5-VL, the file path was passed and loaded internally by qwen_vl_utils.process_vision_info().

Generation used max_new_tokens = 512 and greedy decoding without sampling (do_sample = False, or sampling = False with num_beams = 1 for MiniCPM-V-4.5); other generation settings were library defaults unless specified in the table.

Offline mode was enforced with HF_HUB_OFFLINE = 1 and TRANSFORMERS_OFFLINE = 1.

Hardware: single NVIDIA Tesla V100-SXM2 GPU (32 GB), CUDA 12.2, Python 3.10; flash-attn was not used.

Transformers versions and numerical precision for each model are listed in **Supplementary Table S2A**.

[1]Remote code: model-specific code distributed with the model repository and executed with trust_remote_code = True.

**Supplementary Table S3. Responses of the vision-language models to synthetic control images without anatomical content**

| Model | Black | Gray | Noise | Positive responses |
|---|---|---|---|---|
| InternVL3-8B | Yes | Yes | Yes | 3/3 |
| Qwen2.5-VL-7B-Instruct | Yes | Yes | Yes | 3/3 |
| MiniCPM-V-4.5 | No | No | No | 0/3 |
| DeepSeek-VL2-tiny | Yes | Yes | Yes | 3/3 |
| Phi-3.5-vision-instruct | No | No | Yes | 1/3 |
| MedGemma-4B-it | Yes | Yes | No | 2/3 |
| LLaVA-Med v1.5 | Yes | Yes | Yes | 3/3 |
| HuatuoGPT-Vision-7B | Yes | Yes | No | 2/3 |
| Total | 6/8 | 6/8 | 5/8 | 17/24 |

**Supplementary Table S4. Pairwise comparisons of balanced accuracy among all evaluable vision-language models**

| VLM comparison | Balanced accuracy difference, percentage points [95% CI] | FDR-adjusted P value |
|---|---|---|
| MiniCPM-V-4.5 vs LLaVA-Med v1.5 | 30.5 [25.0–35.5] | <.001 |
| MiniCPM-V-4.5 vs DeepSeek-VL2-tiny | 30.5 [25.0–35.5] | <.001 |
| MiniCPM-V-4.5 vs MedGemma-4B-it | 29.5 [24.0–34.5] | <.001 |
| MiniCPM-V-4.5 vs Phi-3.5-vision-instruct | 29.5 [23.5–35.0] | <.001 |
| MiniCPM-V-4.5 vs InternVL3-8B | 29.0 [23.5–34.0] | <.001 |
| MiniCPM-V-4.5 vs HuatuoGPT-Vision-7B | 21.0 [15.0–27.0] | <.001 |
| MiniCPM-V-4.5 vs Qwen2.5-VL-7B-Instruct | 19.5 [13.0–25.5] | <.001 |
| Qwen2.5-VL-7B-Instruct vs LLaVA-Med v1.5 | 11.0 [6.0–16.0] | <.001 |
| Qwen2.5-VL-7B-Instruct vs DeepSeek-VL2-tiny | 11.0 [6.0–16.0] | <.001 |
| Qwen2.5-VL-7B-Instruct vs MedGemma-4B-it | 10.0 [5.0–15.0] | <.001 |
| Qwen2.5-VL-7B-Instruct vs Phi-3.5-vision-instruct | 10.0 [5.0–15.0] | <.001 |
| HuatuoGPT-Vision-7B vs LLaVA-Med v1.5 | 9.5 [5.5–14.0] | <.001 |
| HuatuoGPT-Vision-7B vs DeepSeek-VL2-tiny | 9.5 [5.5–14.0] | <.001 |
| Qwen2.5-VL-7B-Instruct vs InternVL3-8B | 9.5 [4.5–14.5] | <.001 |
| HuatuoGPT-Vision-7B vs MedGemma-4B-it | 8.5 [4.5–13.0] | <.001 |
| HuatuoGPT-Vision-7B vs Phi-3.5-vision-instruct | 8.5 [4.5–13.0] | <.001 |
| HuatuoGPT-Vision-7B vs InternVL3-8B | 8.0 [4.0–12.0] | <.001 |
| Qwen2.5-VL-7B-Instruct vs HuatuoGPT-Vision-7B | 1.5 [-4.0 to 7.0] | 0.842 |
| InternVL3-8B vs LLaVA-Med v1.5 | 1.5 [0.0–3.5] | 0.368 |
| InternVL3-8B vs DeepSeek-VL2-tiny | 1.5 [0.0–3.5] | 0.368 |
| MedGemma-4B-it vs LLaVA-Med v1.5 | 1.0 [0.0–2.5] | 0.609 |
| Phi-3.5-vision-instruct vs LLaVA-Med v1.5 | 1.0 [0.0–2.5] | 0.609 |
| MedGemma-4B-it vs DeepSeek-VL2-tiny | 1.0 [0.0–2.5] | 0.609 |
| Phi-3.5-vision-instruct vs DeepSeek-VL2-tiny | 1.0 [0.0–2.5] | 0.609 |
| InternVL3-8B vs MedGemma-4B-it | 0.5 [-1.0 to 2.5] | 1.000 |
| InternVL3-8B vs Phi-3.5-vision-instruct | 0.5 [-1.5 to 2.5] | 1.000 |

| | | |
|---|---|---|
| LLaVA-Med v1.5 vs DeepSeek-VL2-tiny | 0.0 [0.0–0.0] | 1.000 |
| MedGemma-4B-it vs Phi-3.5-vision-instruct | 0.0 [-2.0 to 2.0] | 1.000 |

Confidence intervals were estimated with a patient-level cluster bootstrap (10,000 resamples). P values were obtained with paired patient-level sign-flip permutation tests (50,000 permutations) and adjusted with the Benjamini–Hochberg method for 28 comparisons; balanced accuracy differed across the eight evaluable models (Friedman test, P < .001). PaliGemma2-3B-pt-896 was not evaluable.

**Supplementary Table S5. Specificity of lesion-negative images according to anatomical level**

| Model | Basal ganglia | Centrum semiovale | Middle cerebellar peduncle | P value (FDR-adjusted) |
|---|---|---|---|---|
| InternVL3-8B | 0/34, 0.0% [0.0–10.2] | 3/32, 9.4% [3.2–24.2] | 0/34, 0.0% [0.0–10.2] | 0.046 |
| Qwen2.5-VL-7B-Instruct | 6/34, 17.6% [8.3–33.5] | 24/32, 75.0% [57.9–86.7] | 0/34, 0.0% [0.0–10.2] | <0.001 |
| MiniCPM-V-4.5 | 26/34, 76.5% [60.0–87.6] | 32/32, 100.0% [89.3–100.0] | 24/34, 70.6% [53.8–83.2] | 0.003 |
| DeepSeek-VL2-tiny | 0/34, 0.0% [0.0–10.2] | 0/32, 0.0% [0.0–10.7] | 0/34, 0.0% [0.0–10.2] | NA |
| Phi-3.5-vision-instruct | 0/34, 0.0% [0.0–10.2] | 2/32, 6.2% [1.7–20.1] | 0/34, 0.0% [0.0–10.2] | 0.100 |
| MedGemma-4B-it | 0/34, 0.0% [0.0–10.2] | 2/32, 6.2% [1.7–20.1] | 0/34, 0.0% [0.0–10.2] | 0.100 |
| LLaVA-Med v1.5 | 0/34, 0.0% [0.0–10.2] | 0/32, 0.0% [0.0–10.7] | 0/34, 0.0% [0.0–10.2] | NA |
| HuatuoGPT-Vision-7B | 5/34, 14.7% [6.4–30.1] | 16/32, 50.0% [33.6–66.4] | 0/34, 0.0% [0.0–10.2] | <0.001 |

Data are the number of true-negative slices/total (specificity, %) [95% CI; Wilson method]. P values compare specificity across the three levels using the Fisher exact test with Benjamini–Hochberg correction.
NA, Not assessable

**Supplementary Table S6. Lesion-detection performance of the vision-language models with the revised prompt (post hoc analysis)**

| VLM | Sensitivity, % [95% CI] | Specificity, % [95% CI] | Balanced accuracy, % [95% CI] | Predicted positive rate, % | Balanced accuracy of original prompt, % |
|---|---|---|---|---|---|
| **General-purpose VLMs** | | | | | |
| InternVL3-8B | 89.0 [81.4–93.7] | 48.0 [38.5–57.7] | 68.5 [63.0–74.0] | 70.5 | 51.5 |
| Qwen2.5-VL-7B-Instruct | 56.0 [46.2–65.3] | 74.0 [64.6–81.6] | 65.0 [58.0–71.5] | 41.0 | 61.0 |
| MiniCPM-V-4.5 | 76.0 [66.8–83.3] | 89.0 [81.4–93.7] | 82.5 [77.0–87.5] | 43.5 | 80.5 |
| DeepSeek-VL2-tiny | 100.0 [96.3–100.0] | 0.0 [0.0–3.7] | 50.0 [50.0–50.0] | 100.0 | 50.0 |
| Phi-3.5-vision-instruct | 0.0 [0.0–3.7] | 100.0 [96.3–100.0] | 50.0 [50.0–50.0] | 0.0 | 51.0 |
| **Medical-purpose VLMs** | | | | | |
| MedGemma-4B-it | 100.0 [96.3–100.0] | 7.0 [3.4–13.7] | 53.5 [51.0–56.0] | 96.5 | 51.0 |
| HuatuoGPT-Vision-7B | 87.0 [79.0–92.2] | 67.0 [57.3–75.4] | 77.0 [71.0–82.5] | 60.0 | 59.5 |

Data are based on 100 lesion-positive and 100 lesion-negative key images from 100 patients. Sensitivity and specificity are given with Wilson 95% confidence intervals; the 95% confidence interval of balanced accuracy was estimated with a patient-level cluster bootstrap (10,000 resamples).
The revised prompt specified the answer format for each item and omitted the vasogenic edema item. LLaVA-Med v1.5 returned no output for any image with the revised prompt, and PaliGemma2-3B-pt-896 returned no structured response; both are not evaluable. The last column repeats the balanced accuracy from Table 2 for comparison; the original prompt remained the primary analysis.

**Supplementary Table S7. Pairwise comparisons of balanced accuracy with the revised prompt (post hoc analysis)**

| VLM comparison | Balanced accuracy difference, percentage points [95% CI] | FDR-adjusted P value |
|---|---|---|
| MiniCPM-V-4.5 vs DeepSeek-VL2-tiny | 32.5 [27.0–37.5] | <.001 |
| MiniCPM-V-4.5 vs Phi-3.5-vision-instruct | 32.5 [27.0–37.5] | <.001 |
| MiniCPM-V-4.5 vs MedGemma-4B-it | 29.0 [23.5–34.0] | <.001 |
| HuatuoGPT-Vision-7B vs DeepSeek-VL2-tiny | 27.0 [21.0–32.5] | <.001 |
| HuatuoGPT-Vision-7B vs Phi-3.5-vision-instruct | 27.0 [21.0–32.5] | <.001 |
| HuatuoGPT-Vision-7B vs MedGemma-4B-it | 23.5 [17.5–29.0] | <.001 |
| InternVL3-8B vs DeepSeek-VL2-tiny | 18.5 [13.0–24.0] | <.001 |
| InternVL3-8B vs Phi-3.5-vision-instruct | 18.5 [13.0–24.0] | <.001 |
| MiniCPM-V-4.5 vs Qwen2.5-VL-7B-Instruct | 17.5 [10.0–25.0] | <.001 |
| InternVL3-8B vs MedGemma-4B-it | 15.0 [9.5–20.0] | <.001 |
| Qwen2.5-VL-7B-Instruct vs DeepSeek-VL2-tiny | 15.0 [8.0–21.5] | <.001 |
| Qwen2.5-VL-7B-Instruct vs Phi-3.5-vision-instruct | 15.0 [8.0–21.5] | <.001 |
| MiniCPM-V-4.5 vs InternVL3-8B | 14.0 [8.5–19.5] | <.001 |
| HuatuoGPT-Vision-7B vs Qwen2.5-VL-7B-Instruct | 12.0 [6.0–18.0] | <.001 |
| Qwen2.5-VL-7B-Instruct vs MedGemma-4B-it | 11.5 [4.5–18.0] | 0.004 |
| HuatuoGPT-Vision-7B vs InternVL3-8B | 8.5 [3.5–13.5] | 0.004 |
| MedGemma-4B-it vs DeepSeek-VL2-tiny | 3.5 [1.0–6.0] | 0.018 |
| MedGemma-4B-it vs Phi-3.5-vision-instruct | 3.5 [1.0–6.0] | 0.018 |
| MiniCPM-V-4.5 vs HuatuoGPT-Vision-7B | 5.5 [-1.0 to 12.0] | 0.141 |
| InternVL3-8B vs Qwen2.5-VL-7B-Instruct | 3.5 [-3.0 to 10.0] | 0.376 |
| DeepSeek-VL2-tiny vs Phi-3.5-vision-instruct | 0.0 [0.0–0.0] | 1.000 |

Differences are the balanced accuracy of the model listed first minus that of the second (the model with the higher balanced accuracy is listed first). Confidence intervals were estimated with a patient-level cluster bootstrap (10,000 resamples). P values were obtained with paired patient-level sign-flip

permutation tests (50,000 permutations) and adjusted with the Benjamini–Hochberg method for 21 comparisons; balanced accuracy differed across the seven evaluable models (Friedman test, $P < .001$). LLaVA-Med v1.5 and PaliGemma2-3B-pt-896 were not evaluable with the revised prompt.
The interval of 0.0 [0.0–0.0] for DeepSeek-VL2-tiny vs. Phi-3.5-vision-instruct reflects that each model gave a single answer for all images

**Supplementary Table S8. Location of the largest enhancing lesion on the key image**

| Location | n (%) |
|---|---|
| **Supratentorial** | 75 (75.0) |
| Frontal lobe | 39 (39.0) |
| Parietal lobe | 11 (11.0) |
| Temporal lobe | 14 (14.0) |
| Occipital lobe | 5 (5.0) |
| Deep gray matter | 6 (6.0) |
| **Infratentorial** | 25 (25.0) |
| Cerebellum | 23 (23.0) |
| Brainstem | 2 (2.0) |

Deep gray matter includes the basal ganglia and thalamus; brainstem includes the pons.

**Supplementary Table S9. Sensitivity, specificity, and balanced accuracy of vision-language models for binary imaging features among lesion-positive key images (n=100)**

**Lesion count (single vs. multiple): positive class = multiple lesions (n=38); negative class = single lesion (n=62)**

| Model | TP | FN | TN | FP | Sensitivity | Specificity | Balanced accuracy |
|---|---|---|---|---|---|---|---|
| InternVL3-8B | 9 | 29 | 62 | 0 | 23.7 [13.0–39.2] | 100.0 [94.2–100.0] | 61.8 [55.3–69.2] |
| Qwen2.5-VL-7B-Instruct | 6 | 32 | 55 | 7 | 15.8 [7.4–30.4] | 88.7 [78.5–94.4] | 52.2 [45.6–59.5] |
| MiniCPM-V-4.5 | 16 | 22 | 47 | 15 | 42.1 [27.9–57.8] | 75.8 [63.8–84.8] | 59.0 [49.0–68.3] |
| DeepSeek-VL2-tiny | 8 | 30 | 62 | 0 | 21.1 [11.1–36.3] | 100.0 [94.2–100.0] | 60.5 [54.3–67.2] |
| Phi-3.5-vision-instruct | 0 | 38 | 62 | 0 | 0.0 [0.0–9.2] | 100.0 [94.2–100.0] | 50.0 [50.0–50.0] |
| MedGemma-4B-it | 20 | 18 | 56 | 6 | 52.6 [37.3–67.5] | 90.3 [80.5–95.5] | 71.5 [63.0–80.0] |
| LLaVA-Med v1.5 | 36 | 2 | 14 | 48 | 94.7 [82.7–98.5] | 22.6 [14.0–34.4] | 58.7 [52.5–65.2] |
| HuatuoGPT-Vision-7B | 18 | 20 | 60 | 2 | 47.4 [32.5–62.7] | 96.8 [89.0–99.1] | 72.1 [63.7–80.3] |

**Location (supra- vs. infratentorial): positive class = infratentorial (n=25); negative class = supratentorial (n=75)**

| Model | TP | FN | TN | FP | Sensitivity | Specificity | Balanced accuracy |
|---|---|---|---|---|---|---|---|
| InternVL3-8B | 16 | 9 | 72 | 3 | 64.0 [44.5–79.8] | 96.0 [88.9–98.6] | 80.0 [70.0–89.3] |
| Qwen2.5-VL-7B-Instruct | 14 | 11 | 72 | 3 | 56.0 [37.1–73.3] | 96.0 [88.9–98.6] | 76.0 [65.4–86.2] |
| MiniCPM-V-4.5 | 20 | 5 | 52 | 23 | 80.0 [60.9–91.1] | 69.3 [58.2–78.6] | 74.7 [64.4–84.0] |
| DeepSeek-VL2-tiny | 0 | 25 | 0 | 75 | 0.0 [0.0–13.3] | 0.0 [0.0–4.9] | 0.0 [0.0–0.0] |
| Phi-3.5-vision-instruct | 25 | 0 | 39 | 36 | 100.0 [86.7–100.0] | 52.0 [40.9–62.9] | 76.0 [70.5–81.5] |
| MedGemma-4B-it | 13 | 12 | 66 | 9 | 52.0 [33.5–70.0] | 88.0 [78.7–93.6] | 70.0 [59.3–80.6] |
| LLaVA-Med v1.5 | 8 | 17 | 75 | 0 | 32.0 [17.2–51.6] | 100.0 [95.1–100.0] | 66.0 [57.1–75.0] |
| HuatuoGPT-Vision-7B | 23 | 2 | 71 | 4 | 92.0 [75.0–97.8] | 94.7 [87.1–97.9] | 93.3 [86.5–98.4] |

**Enhancement (ring vs. non-ring): positive class = ring enhancement (n=21); negative class = non-ring enhancement (n=79)**

| Model | TP | FN | TN | FP | Sensitivity | Specificity | Balanced accuracy |
|---|---|---|---|---|---|---|---|
| InternVL3-8B | 21 | 0 | 13 | 66 | 100.0 [84.5–100.0] | 16.5 [9.9–26.1] | 58.2 [54.3–62.6] |
| Qwen2.5-VL-7B-Instruct | 21 | 0 | 0 | 79 | 100.0 [84.5–100.0] | 0.0 [0.0–4.6] | 50.0 [50.0–50.0] |

| | | | | | | | |
|---|---|---|---|---|---|---|---|
| MiniCPM-V-4.5 | 20 | 1 | 16 | 63 | 95.2 [77.3–99.2] | 20.3 [12.9–30.4] | 57.7 [50.7–63.8] |
| DeepSeek-VL2-tiny | 0 | 21 | 0 | 79 | 0.0 [0.0–15.5] | 0.0 [0.0–4.6] | 0.0 [0.0–0.0] |
| Phi-3.5-vision-instruct | 0 | 21 | 79 | 0 | 0.0 [0.0–15.5] | 100.0 [95.4–100.0] | 50.0 [50.0–50.0] |
| MedGemma-4B-it | 16 | 5 | 41 | 38 | 76.2 [54.9–89.4] | 51.9 [41.1–62.6] | 64.0 [53.3–74.4] |
| LLaVA-Med v1.5 | 17 | 4 | 25 | 54 | 81.0 [60.0–92.3] | 31.6 [22.4–42.5] | 56.3 [45.6–65.7] |
| HuatuoGPT-Vision-7B | 21 | 0 | 5 | 74 | 100.0 [84.5–100.0] | 6.3 [2.7–14.0] | 53.2 [50.6–56.1] |

**Necrosis: positive class = necrosis present (n=49); negative class = necrosis absent (n=51)**

| Model | TP | FN | TN | FP | Sensitivity | Specificity | Balanced accuracy |
|---|---|---|---|---|---|---|---|
| InternVL3-8B | 7 | 42 | 49 | 2 | 14.3 [7.1–26.7] | 96.1 [86.8–98.9] | 55.2 [49.8–60.9] |
| Qwen2.5-VL-7B-Instruct | 49 | 0 | 8 | 43 | 100.0 [92.7–100.0] | 15.7 [8.2–28.0] | 57.8 [53.2–63.0] |
| MiniCPM-V-4.5 | 43 | 6 | 6 | 45 | 87.8 [75.8–94.3] | 11.8 [5.5–23.4] | 49.8 [43.5–56.1] |
| DeepSeek-VL2-tiny | 0 | 49 | 0 | 51 | 0.0 [0.0–7.3] | 0.0 [0.0–7.0] | 0.0 [0.0–0.0] |
| Phi-3.5-vision-instruct | 47 | 2 | 1 | 50 | 95.9 [86.3–98.9] | 2.0 [0.3–10.3] | 48.9 [45.4–52.3] |
| MedGemma-4B-it | 25 | 24 | 43 | 8 | 51.0 [37.5–64.4] | 84.3 [72.0–91.8] | 67.7 [58.9–76.1] |
| LLaVA-Med v1.5 | 49 | 0 | 0 | 51 | 100.0 [92.7–100.0] | 0.0 [0.0–7.0] | 50.0 [50.0–50.0] |
| HuatuoGPT-Vision-7B | 48 | 1 | 4 | 47 | 98.0 [89.3–99.6] | 7.8 [3.1–18.5] | 52.9 [49.0–57.1] |

**Mass effect or midline shift: positive class = mass effect present (n=13); negative class = mass effect absent (n=87)**

| Model | TP | FN | TN | FP | Sensitivity | Specificity | Balanced accuracy |
|---|---|---|---|---|---|---|---|
| InternVL3-8B | 0 | 13 | 87 | 0 | 0.0 [0.0–22.8] | 100.0 [95.8–100.0] | 50.0 [50.0–50.0] |
| Qwen2.5-VL-7B-Instruct | 11 | 2 | 28 | 59 | 84.6 [57.8–95.7] | 32.2 [23.3–42.6] | 58.4 [46.5–69.0] |
| MiniCPM-V-4.5 | 12 | 1 | 51 | 36 | 92.3 [66.7–98.6] | 58.6 [48.1–68.4] | 75.5 [64.4–82.9] |
| DeepSeek-VL2-tiny | 0 | 13 | 0 | 87 | 0.0 [0.0–22.8] | 0.0 [0.0–4.2] | 0.0 [0.0–0.0] |
| Phi-3.5-vision-instruct | 0 | 13 | 87 | 0 | 0.0 [0.0–22.8] | 100.0 [95.8–100.0] | 50.0 [50.0–50.0] |
| MedGemma-4B-it | 11 | 2 | 74 | 13 | 84.6 [57.8–95.7] | 85.1 [76.1–91.1] | 84.8 [72.8–94.3] |
| LLaVA-Med v1.5 | 0 | 13 | 87 | 0 | 0.0 [0.0–22.8] | 100.0 [95.8–100.0] | 50.0 [50.0–50.0] |
| HuatuoGPT-Vision-7B | 11 | 2 | 76 | 11 | 84.6 [57.8–95.7] | 87.4 [78.8–92.8] | 86.0 [74.1–95.2] |

TP, true positive; FN, false negative; TN, true negative; FP, false positive. Images in which the model did not report a lesion and invalid responses (blank, not applicable, repeated item names, or outside the answer categories) were counted as incorrect, i.e., as FN when the reference was the positive class and as FP when it was the negative class.
Sensitivity and specificity are given with Wilson 95% confidence intervals; balanced accuracy is the mean of sensitivity and specificity, with a 95% confidence interval from a percentile bootstrap with 2,000 resamples. A balanced accuracy of 50.0% with an interval of 50.0–50.0 indicates that all images were assigned to a single class (invalid responses counted as incorrect).
For DeepSeek-VL2-tiny, all responses to these items were invalid and were counted as incorrect; its sensitivity, specificity, and balanced accuracy of 0% reflect invalid responses, not discrimination.

**Supplementary Table S10. Accuracy of exact lesion count (1, 2, 3, 4, 5, or ≥6 lesions) among lesion-positive key images (n=100)**

| Model | Correct/total | Accuracy, % [95% CI] | No lesion reported, n | Macro-F1 |
|---|---|---|---|---|
| Majority-class baseline ("1") | 62/100 | 62.0 | | 0.13 |
| InternVL3-8B | 64/100 | 64.0 [54.2–72.7] | 0 | 0.18 |
| Qwen2.5-VL-7B-Instruct | 55/100 | 55.0 [45.2–64.4] | 8 | 0.12 |
| MiniCPM-V-4.5 | 53/100 | 53.0 [43.3–62.5] | 21 | 0.26 |
| DeepSeek-VL2-tiny | 65/100 | 65.0 [55.3–73.6] | 0 | 0.18 |
| Phi-3.5-vision-instruct | 62/100 | 62.0 [52.2–70.9] | 0 | 0.13 |
| MedGemma-4B-it | 66/100 | 66.0 [56.3–74.5] | 0 | 0.28 |
| LLaVA-Med v1.5 | 27/100 | 27.0 [19.3–36.4] | 0 | 0.10 |
| HuatuoGPT-Vision-7B | 64/100 | 64.0 [54.2–72.7] | 2 | 0.22 |

Reference distribution: 1 lesion, n=62; 2, n=14; 3, n=6; 4, n=4; 5, n=3; ≥6, n=11. Accuracy is the proportion of images for which the model gave the correct category; images without a reported lesion and invalid responses were counted as incorrect. Macro-F1 is the unweighted mean of the F1 scores of the six reference categories; an always-"1" classifier has a macro-F1 of 0.13. The 95% confidence intervals were calculated with the Wilson method. The 95% confidence interval included the baseline accuracy (62%) in all models except LLaVA-Med v1.5, whose accuracy was lower than the baseline; the lower bound of the confidence interval did not exceed the baseline in any model.

### Supplementary Table S11. Imaging feature assessment conditional on lesion detection and stratified by single- vs. multiple-lesion cases

**A. Conditional on detection: lesion-positive key images in which the model reported a lesion**

| Model | Images with a lesion reported, n | Lesion count (single vs. multiple) | | Lesion count (exact) | Laterality | Location | |
|---|---|---|---|---|---|---|---|
| | | Accuracy, % | Balanced accuracy, % | Accuracy, % | Accuracy, % | Accuracy, % | Balanced accuracy, % |
| Qwen2.5-VL-7B-Instruct | 92 | 66.3 [56.2–75.1] | 58.1 [52.6–64.7] | 59.8 [49.6–69.2] | 44.6 [34.8–54.7] | 84.8 [76.1–90.7] | 75.8 [66.1–85.7] |
| MiniCPM-V-4.5 | 79 | 79.7 [69.6–87.1] | 74.8 [65.8–84.2] | 67.1 [56.1–76.4] | 50.6 [39.8–61.4] | 91.1 [82.8–95.6] | 92.4 [85.6–97.5] |
| HuatuoGPT-Vision-7B | 98 | 79.6 [70.6–86.4] | 73.7 [65.7–81.4] | 65.3 [55.5–74.0] | 80.6 [71.7–87.2] | 95.9 [90.0–98.4] | 94.6 [88.2–99.4] |

| Model (continued) | Images with a lesion reported, n | Enhancement pattern | | Necrosis | | Mass effect or midline shift | |
|---|---|---|---|---|---|---|---|
| | | Accuracy, % | Balanced accuracy, % | Accuracy, % | Balanced accuracy, % | Accuracy, % | Balanced accuracy, % |
| Qwen2.5-VL-7B-Instruct | 92 | 22.8 [15.4–32.4] | 50.0 [50.0–50.0] | 53.3 [43.1–63.1] | 50.0 [50.0–50.0] | 33.7 [24.9–43.8] | 55.0 [43.0–65.0] |
| MiniCPM-V-4.5 | 79 | 45.6 [35.0–56.5] | 63.6 [57.9–69.4] | 62.0 [51.0–71.9] | 55.1 [47.6–63.0] | 79.7 [69.6–87.1] | 84.8 [74.9–92.4] |
| HuatuoGPT-Vision-7B | 98 | 26.5 [18.8–36.0] | 53.2 [50.6–56.2] | 53.1 [43.3–62.6] | 53.1 [49.0–57.4] | 88.8 [81.0–93.6] | 87.0 [75.8–96.3] |

Models that reported a lesion in all 100 lesion-positive images (InternVL3-8B, DeepSeek-VL2-tiny, Phi-3.5-vision-instruct, MedGemma-4B-it, LLaVA-Med v1.5) are not shown because their results are identical to the end-to-end results in Tables 4 and 5 and Supplementary Tables S9 and S10. Invalid responses were counted as incorrect. Accuracy is given with the Wilson 95% confidence interval; balanced accuracy is the mean of sensitivity and specificity, with a 95% confidence interval from a percentile bootstrap with 2,000 image resamples. Positive classes: multiple lesions, infratentorial location, ring enhancement, necrosis, and mass effect or midline shift. Balanced accuracy was not calculated for laterality (three categories) or exact lesion count (six categories).

**B. Single-lesion cases (n=62)**

| Model | Laterality | Location | | Enhancement pattern | | Necrosis | | Mass effect or midline shift | |
|---|---|---|---|---|---|---|---|---|---|
| | Accuracy, % | Accuracy, % | Balanced accuracy, % | Accuracy, % | Balanced accuracy, % | Accuracy, % | Balanced accuracy, % | Accuracy, % | Balanced accuracy, % |
| Majority-class baseline | 59.7 | 75.8 | — | 87.1 | — | 58.1 | — | 87.1 | — |
| InternVL3-8B | 32.3 [22.0–44.6] | 88.7 [78.5–94.4] | 78.9 [65.6–91.3] | 27.4 [17.9–39.6] | 58.3 [53.6–63.5] | 59.7 [47.3–71.0] | 53.0 [46.0–60.6] | 87.1 [76.6–93.3] | 50.0 [50.0–50.0] |
| Qwen2.5-VL-7B-Instruct | 50.0 [37.9–62.1] | 87.1 [76.6–93.3] | 75.6 [63.0–88.9] | 12.9 [6.7–23.4] | 50.0 [50.0–50.0] | 53.2 [41.0–65.1] | 59.7 [53.8–66.7] | 35.5 [24.7–47.9] | 57.6 [42.2–68.3] |
| MiniCPM-V-4.5 | 45.2 [33.4–57.5] | 75.8 [63.8–84.8] | 79.5 [68.9–89.4] | 33.9 [23.3–46.3] | 62.0 [56.6–67.9] | 41.9 [30.5–54.3] | 47.9 [38.6–56.1] | 67.7 [55.4–78.0] | 76.2 [60.9–87.1] |
| DeepSeek-VL2-tiny | 59.7 [47.3–71.0] | 0.0 [0.0–5.8]† | 0.0 [0.0–0.0]† | 0.0 [0.0–5.8]† | 0.0 [0.0–0.0]† | 0.0 [0.0–5.8]† | 0.0 [0.0–0.0]† | 0.0 [0.0–5.8]† | 0.0 [0.0–0.0]† |
| Phi-3.5-vision-instruct | 59.7 [47.3–71.0] | 71.0 [58.7–80.8] | 80.9 [73.8–87.5] | 87.1 [76.6–93.3] | 50.0 [50.0–50.0] | 41.9 [30.5–54.3] | 49.5 [44.4–53.8] | 87.1 [76.6–93.3] | 50.0 [50.0–50.0] |
| MedGemma-4B-it | 69.4 [57.0–79.4] | 82.3 [71.0–89.8] | 67.9 [55.0–81.0] | 58.1 [45.7–69.5] | 75.9 [69.3–82.4] | 75.8 [63.8–84.8] | 73.3 [62.3–84.3] | 83.9 [72.8–91.0] | 85.4 [69.9–95.4] |
| LLaVA-Med v1.5 | 22.6 [14.0–34.4] | 83.9 [72.8–91.0] | 66.7 [55.0–79.4] | 37.1 [26.2–49.5] | 58.6 [43.7–69.4] | 41.9 [30.5–54.3] | 50.0 [50.0–50.0] | 87.1 [76.6–93.3] | 50.0 [50.0–50.0] |
| HuatuoGPT-Vision-7B | 91.9 [82.5–96.5] | 91.9 [82.5–96.5] | 92.4 [84.2–98.9] | 21.0 [12.7–32.6] | 54.6 [50.9–59.1] | 46.8 [34.9–59.0] | 53.6 [47.5–60.3] | 91.9 [82.5–96.5] | 90.0 [74.9–99.0] |

Majority-class baselines were calculated within each stratum and are shown for reference (not statistically tested).

†All responses for this item were invalid (item names repeated, or "left"/"right" instead of an anatomical location). Invalid responses were counted as incorrect, so accuracy and balanced accuracy are 0% and reflect invalid responses rather than discrimination.

**C. Multiple-lesion cases (n=38)**

| Model | Laterality | Location | | Enhancement pattern | | Necrosis | | Mass effect or midline shift | |
|---|---|---|---|---|---|---|---|---|---|
| | Accuracy, % | Accuracy, % | Balanced accuracy, % | Accuracy, % | Balanced accuracy, % | Accuracy, % | Balanced accuracy, % | Accuracy, % | Balanced accuracy, % |
| Majority-class baseline | 76.3 | 73.7 | — | 65.8 | — | 60.5 | — | 86.8 | — |
| InternVL3-8B | 26.3 [15.0–42.0] | 86.8 [72.7–94.2] | 81.4 [65.0–96.4] | 44.7 [30.1–60.3] | 58.0 [51.9–65.4] | 50.0 [34.8–65.2] | 58.7 [52.1–67.4] | 86.8 [72.7–94.2] | 50.0 [50.0–50.0] |
| Qwen2.5-VL-7B-Instruct | 28.9 [17.0–44.8] | 84.2 [69.6–92.6] | 76.4 [59.9–92.9] | 34.2 [21.2–50.1] | 50.0 [50.0–50.0] | 63.2 [47.3–76.6] | 53.3 [50.0–61.1] | 44.7 [30.1–60.3] | 59.7 [36.7–76.5] |
| MiniCPM-V-4.5 | 31.6 [19.1–47.5] | 65.8 [49.9–78.8] | 67.1 [48.8–83.8] | 39.5 [25.6–55.3] | 52.2 [41.0–61.4] | 60.5 [44.7–74.4] | 52.3 [41.7–63.2] | 55.3 [39.7–69.9] | 74.2 [65.2–82.8] |
| DeepSeek-VL2-tiny | 13.2 [5.8–27.3] | 0.0 [0.0–9.2]† | 0.0 [0.0–0.0]† | 0.0 [0.0–9.2]† | 0.0 [0.0–0.0]† | 0.0 [0.0–9.2]† | 0.0 [0.0–0.0]† | 0.0 [0.0–9.2]† | 0.0 [0.0–0.0]† |
| Phi-3.5-vision-instruct | 13.2 [5.8–27.3] | 52.6 [37.3–67.5] | 67.9 [59.5–77.1] | 65.8 [49.9–78.8] | 50.0 [50.0–50.0] | 57.9 [42.2–72.1] | 47.8 [42.9–50.0] | 86.8 [72.7–94.2] | 50.0 [50.0–50.0] |
| MedGemma-4B-it | 55.3 [39.7–69.9] | 73.7 [58.0–85.0] | 72.5 [54.2–88.2] | 55.3 [39.7–69.9] | 56.8 [39.6–73.8] | 55.3 [39.7–69.9] | 58.4 [42.1–73.7] | 86.8 [72.7–94.2] | 83.9 [58.0–97.2] |
| LLaVA-Med v1.5 | 39.5 [25.6–55.3] | 81.6 [66.6–90.8] | 65.0 [50.0–80.8] | 50.0 [34.8–65.2] | 56.5 [40.2–71.0] | 60.5 [44.7–74.4] | 50.0 [50.0–50.0] | 86.8 [72.7–94.2] | 50.0 [50.0–50.0] |
| HuatuoGPT-Vision-7B | 57.9 [42.2–72.1] | 97.4 [86.5–99.5] | 95.0 [83.3–100.0] | 34.2 [21.2–50.1] | 50.0 [50.0–50.0] | 60.5 [44.7–74.4] | 50.0 [50.0–50.0] | 78.9 [63.7–88.9] | 79.4 [53.9–94.3] |

Lesion count items are not shown because they are determined by the stratum. The majority-class baseline is the accuracy of a classifier that always predicts the most frequent reference category within the stratum. Invalid responses were counted as incorrect. Confidence intervals were calculated as in panel A.

†All responses for this item were invalid (item names repeated, or "left"/"right" instead of an anatomical location). Invalid responses were counted as incorrect, so accuracy and balanced accuracy are 0% and reflect invalid responses rather than discrimination.
